# Pan-Cancer Profiling of Mitotic Topology & Mitotic Errors: Insights into Prognosis, Genomic Alterations, and Immune Landscape

**DOI:** 10.1101/2025.06.07.25329181

**Authors:** Mostafa Jahanifar, Muhammad Dawood, Neda Zamanitajeddin, Adam Shephard, Brinder Singh Chohan, Christof A Bertram, Noorul Wahab, Mark Eastwood, Marc Aubreville, Shan E Ahmed Raza, Fayyaz Minhas, Nasir Rajpoot

## Abstract

Mitotic activity and mitotic errors are critical biomarkers of cancer proliferation and progression. While the role of mitosis count in diverse cancer types has been studied, its characterization through organizational patterns in whole slide images remains underexplored within a unified analytical framework. In this study, we present a novel methodology leveraging social network analysis to quantify mitotic topology within the tumor microenvironment, and introduce an atypical mitosis detection model to quantify mitotic errors in histology slides based on the density of atypical mitoses. We extensively investigate the significance of the Pan-Cancer Mitotic Activity Network (PACMAN) for profiling mitotic activity and mitotic errors across various cancers. Our approach is based on deep mining of standard H&E histology images, offering a cost-effective and efficient solution suitable for broad clinical use. Analyzing 9,141 cases from The Cancer Genome Atlas (TCGA) across 31 cancers, we demonstrate that mitotic topological features and atypical mitoses serve as independent prognostic indicators for most cancers. Also, we provide new insights into their association with tumor proliferation, genomic alterations, and immune evasion. Finally, we make available comprehensive resources to facilitate future research in this domain.

## INTRODUCTION

Fundamentally, cancer is a disease of uncontrolled cell division, and at the heart of it, the mitotic cycle serves as an engine and the gateway through which many key oncogenic events unfold. Typically, heightened proliferative signaling drives an increased mitotic activity while defects in cell-cycle checkpoints and DNA repair mechanisms allow mitotic errors to persist and propagate^1^. These aberrations not only contribute to genomic instability^2^, an enabling characteristic of the hallmarks of cancer^2^, but also reinforce the growth, survival, and adaptive advantages that cancer cells depend upon. Therefore, quantifying mitotic activity and mitotic errors is necessary to deepen our understanding of tumor biology and improve clinical outcomes.

Despite its importance, the investigation of mitosis in cancer faces three key challenges. First, studies assessing mitotic activity through histopathology are often limited to specific cancer types, lacking a unified, pan-cancer framework^3^. Second, histopathological reporting typically relies on mitotic count within small hotspots of variable size^3,4^, ignoring the spatial topology of mitoses across the tumor microenvironment (TME). Third, while errors in mitosis can drive aneuploidy and genomic instability^1^, the clinical significance of atypical mitosis remains understudied beyond a few cancers like adrenocortical^5^, breast^6^, urothelial^7^, lung^8^, and pancreatic^9^ carcinomas as well as canine cutaneous mast cell tumors^10^. Fortunately, mitotic errors are morphologically manifesting as atypical mitotic figures (such as multipolar mitoses, lagging chromosomes, and anaphase bridges^6^ as seen in Figure 1C) in standard H&E samples and therefore can be quantified at large scale using deep learning (DL) models.

**Figure 1:**
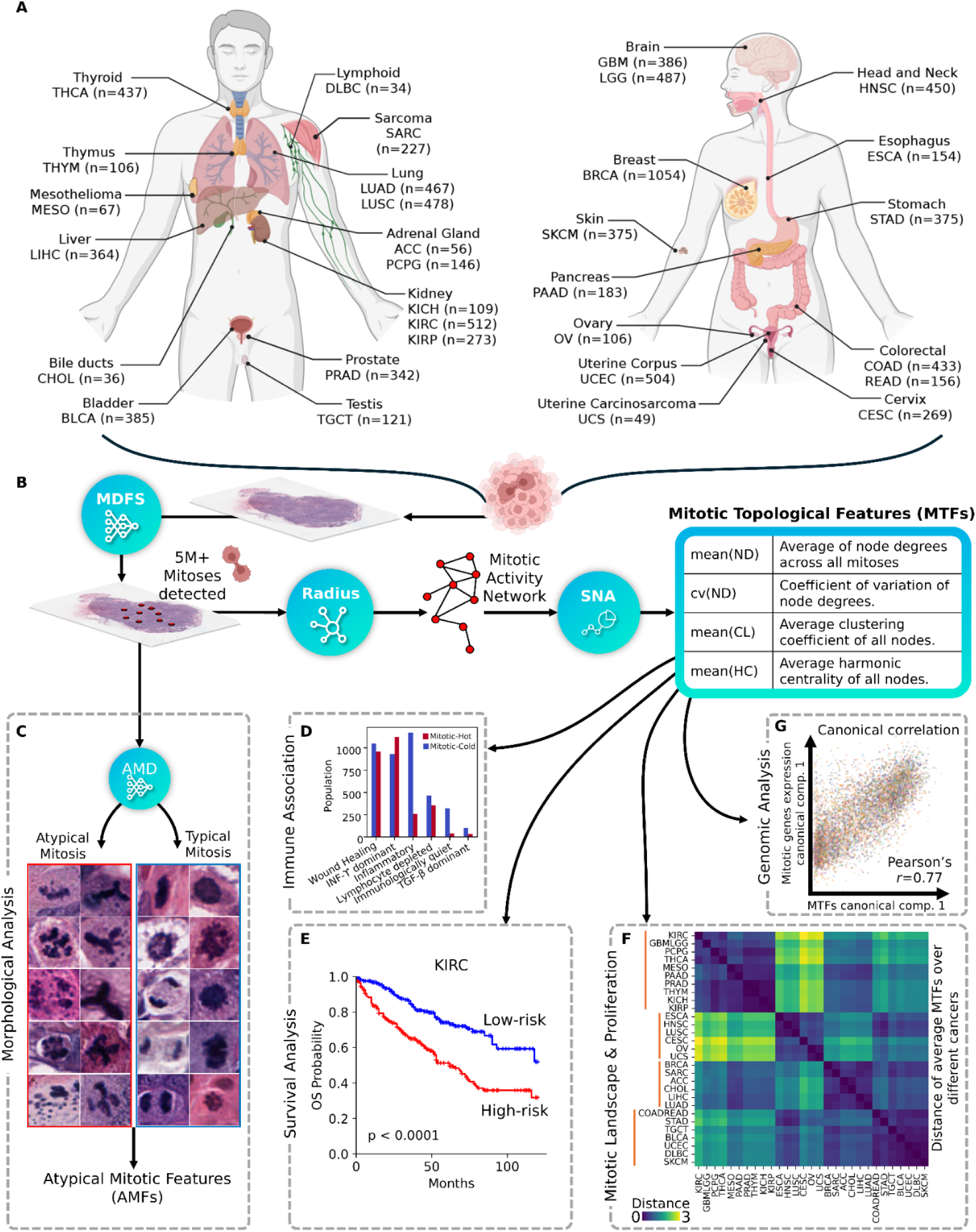
Pan-cancer profiling of mitotic topology and errors. (A) 9141 cases across 31 cancers from TCGA are investigated in this work. TCGA study abbreviations can be found here: https://gdc.cancer.gov/resources-tcga-users/tcga-code-tables/tcga-study-abbreviations. (B) Our proposed pipeline for Mitotic Topological Features (MTFs) extraction. (C) Atypical Mitosis Detection (AMD) model is introduced to further subclassify mitoses into typical and atypical morphologies, which are used to drive Atypical Mitotic Features (AMFs). Leveraging MTFs, we perform pan-cancer analyses that can reveal how mitotic activity is related to (D) immune landscape, (E) patient prognosis, (F) proliferation landscape, and (G) genomics.

To address the gaps above, we leverage digital pathology to systematically analyze mitotic activity and errors across the TME using state-of-the-art DL and graph-based approaches in a unified framework. Our study draws upon a large-scale dataset of 9,141 cases from The Cancer Genome Atlas (TCGA), encompassing 31 diverse tumor types (Figure 1A). We introduce a novel network-based approach for quantifying mitotic activity in whole-slide images (WSIs) of routine H&E stained sections that captures the topology of mitoses (Figure 1B). Furthermore, we systematically study mitotic errors through automatic atypical mitoses detection (Figure 1C), extending our mitotic feature database and enabling the first large-scale analysis of aberrant cell division patterns in situ.

By integrating these features with established clinical^11^, genomic^12,13^, and immunological^14^ data, we perform comprehensive pan-cancer analyses revealing how mitotic activity and errors relate to prognosis, genomic alterations, immune landscape, and proliferation heterogeneity across tumors (Figure 1D-G). Overall, our work not only provides novel methodologies to systematically characterize mitotic behavior in cancer but also serves as a resource for the research community (see “Materials availability” section).

## RESULTS

### Mitotic landscape of cancer

Our method is based on deep mining of H&E slides (Figure 1B), where we first detect mitotic figures using an extensively validated algorithm called “Mitosis Detection Fast and Slow (MDFS)”^15^. Treating each detected mitosis as a network node, we connect nodes based on their distances to create a “Mitotic Activity Network (MAN)”. We then employ social network analysis (SNA) to measure local node properties in the network^16^, including mitotic connectivity (node degree or ND), mitotic clustering (clustering coefficients or CL), and mitotic proximity (harmonic centrality or HC). Statistics of these network measures form a new class of Mitotic Topological Features (MTFs).

Crucially, to systematically study mitotic errors, we develop an Atypical Mitosis Detector (AMD) model that further classifies each detected mitosis into typical or atypical morphologies (Figure 1C). Then, we devise three Atypical Mitotic Features (AMFs) to quantify the atypical mitoses activity in TME: Number of “Atypical Mitoses in an Atypical Hotspot (AMAH)”, Number of “Atypical Mitoses in mitotic Hotspot (AMH)”, and “Atypical mitosis Fraction in WSI (AFW)”. The mitotic feature set for TCGA is provided in Table S1. Further details on the methodology are provided in the Supplemental Methods section.

#### MTFs capture mitotic activity through molecular correlation analyses

Through cross-validated canonical correlation analyses (Figure 1G), we established a strong linear relationship between MTFs and the expression of genes or proteins involved in mitotic cell cycle based on gene ontology (GO: 0000278), achieving an overall high correlation on test sets (Pearson’s *r*=0.77, p < 0.0001). MTFs also correlate strongly with the established Mitotic Network Activity Index (MNAI)^17,18^, as seen in Figure S1A. In particular, mean(CL) exhibited notably high and statistically significant correlations across multiple cancers: BRCA (*r*=0.68), LIHC (*r*=0.62), LUAD (*r*=0.65), OV (*r*=0.62), SARC (*r*=0.68), and when considered pan-cancer (*r*=0.70), with FDR-adjusted p-values (q-values)<0.0001. Other features, such as HSC, mean(ND), and per99(ND), also correlated well with MNAI, supporting the idea that network-based characterization of mitotic distribution is a robust measure of tumor mitotic activity.

To broaden the applicability of MTFs, we investigated their correlation with 16,219 protein-coding genes and identified a subset of 182 genes with absolute correlation values greater than 0.60 and q-values<0.001. This subset, termed the Pan-Cancer Mitotic Activity Network (PACMAN) genes, includes 46 (of 53) MNAI genes and identifies 136 additional genes associated with mitotic activity (Figure 2A and Table S2). PACMAN genes were further validated through gene set enrichment analysis (Figures 1B and S1B) where we found them significantly enriched in pathways, biological processes, cellular components, and molecular functions related to mitosis.

**Figure 2:**
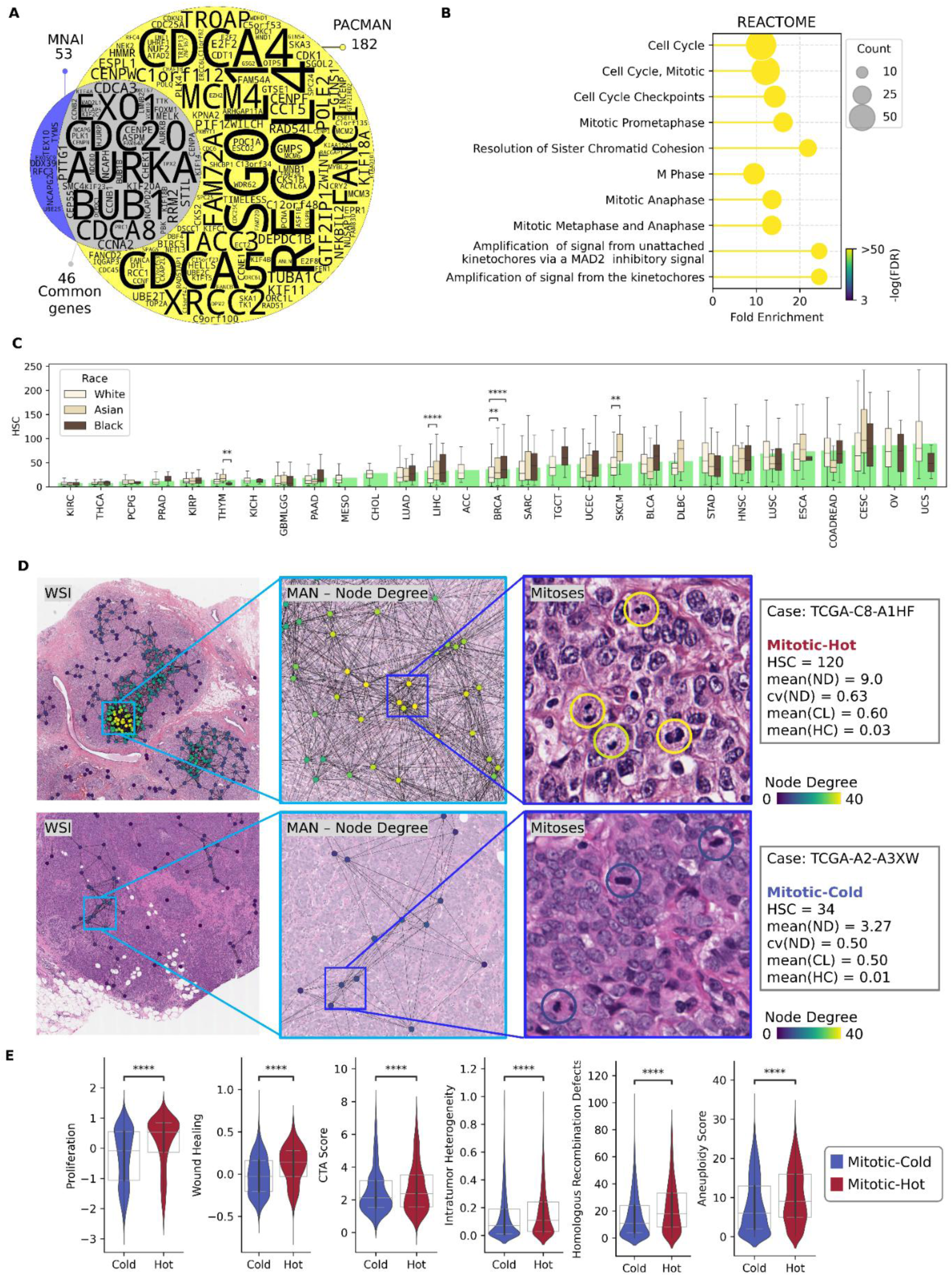
MTFs describe the mitotic landscape of cancer. (A) PACMAN gene set overlap with MNAI^17,18^. (B) Top 10 pathways enriched in gene ontology analyses using PACMAN genes. (C) Distribution of mitotic activity (based on HSC) in different cancers (green bars) and races (box plots). Whiskers indicate the range within 1.5×IQR. (D) Examples of Mitotic-Hot and Mitotic-Cold cases with their corresponding MAN. Nodes are colored based on “Node Degree” measure (more examples at https://tiademos.dcs.warwick.ac.uk/bokeh_app?demo=pacman). (E) Distribution of tumor characteristics across Mitotic-Hot and Mitotic-Cold groups on pan-cancer level.

#### Mitotic activity varies between different cancers and ethnic backgrounds

The distribution of mitoses in TME changes across different cancers as shown in the distance heatmap in Figure 1F, showing Euclidean distances between mean MTFs of different cancer pairs. It is evident that the topology of MAN is very similar in CESC, OV, and UCS but different in UCEC, which shows a similar distribution to DLBC and SKCM. Squamous cell carcinomas (CESC, HNSC, and LUSC) also have similar average mitotic distribution.

In terms of the average mitotic activity (HSC) across different cancers (green bars in Figure 2C), KIRC shows the lowest and UCS the highest mitotic activity. Differences in mitotic activity among different races in each cancer type can also be observed, in line with known differences for BRCA^19^. Moreover, the trends we see in THYM, LIHC, and SKCM point to potential biological disparities, although potential confounding factors warrant cautious interpretation and highlight the need for further investigations in more controlled cohorts.

Furthermore, we clustered the cases into two subgroups of Mitotic-Hot and Mitotic-Cold based on the density of mitosis (HSC) and using Gaussian mixture models in each cancer type (Materials and Methods). Intuitively, Mitotic-Hot tumors exhibit higher mitotic activity, indicating rapid proliferation in comparison to Mitotic-Cold tumors. This stratification enables a more detailed comparison of tumors with high versus low proliferative activity. Examples of Mitotic-Hot and Mitotic-Cold cases are shown in Figure 2D.

Population ratios of Mitotic-Hot to Mitotic-Cold subgroups vary in different cancers (Figure S2A). In most cancers, 20%-30% of cases are Mitotic-Hot except for COADREAD, GBMLGG, and LIHC which have a higher share of this subgroup. On the other hand, MESO and PAAD show less than 20% Mitotic-Hot cases.

On investigating the distribution of MTFs over Mitotic-Hot and Mitotic-Cold subgroups for pan-cancer and cancer-specific scenarios (Figure S5B-C), we find MANs tend to be more connected, clustered, and compact in Mitotic-Hot cases compared to Mitotic-Cold cases. Furthermore, cancers from the female reproductive system (OV, CESC, and UCS), squamous cell carcinoma type (HNSC and LUSC), and upper GI (ESCA and STAD) have relatively higher mitotic activity than other cancers.

#### Tumor characteristics are different between Mitotic-Hot and Mitotic-Cold groups

Figure 2E displays violin plots comparing tumor characteristics between Mitotic-Hot and Mitotic-Cold cases. Expectedly, the Mitotic-Hot group shows significantly higher proliferation rates in pan-cancer and cancer-specific (Figure S3A) scenarios. Proliferation scores, derived from genomic data^14^, are also highly correlated with MTFs (Pearson’s r=0.7, q-value<0.0001 in pan-cancer correlation with mean(CL), Fig. S3B). Similarly, the elevated “Cancer-Testis Antigen (CTA)” score (specifically in BRCA, KIRC, LUAD, and UCEC; Figure S3C) and “Wound Healing” signature levels observed in Mitotic-Hot tumors reflect the heightened proliferative activity of these tumors. CTAs contribute to tumor cellular fitness by promoting cell cycle progression and resisting cell death^20^, while the activation of wound healing pathways mirrors the persistent tissue remodeling and inflammation co-opted by tumors for growth and survival^21^. Furthermore, in line with previous findings^17^, the Mitotic-Hot group shows significantly higher intratumor heterogeneity, homologous recombination defects, and aneuploidy.

### Mitotic topological features are associated with genomic alterations

#### MTFs reveal gene mutations associated with mitotic activity

Associations between gene mutation status (mutated vs. wild-type) and MTF values were evaluated using the AUC metric^22^ ranging from –1 to 1 (Figure 3A, Table S3), where positive values indicate higher mitotic activity in mutated cases and negative values indicate lower activity.

**Figure 3:**
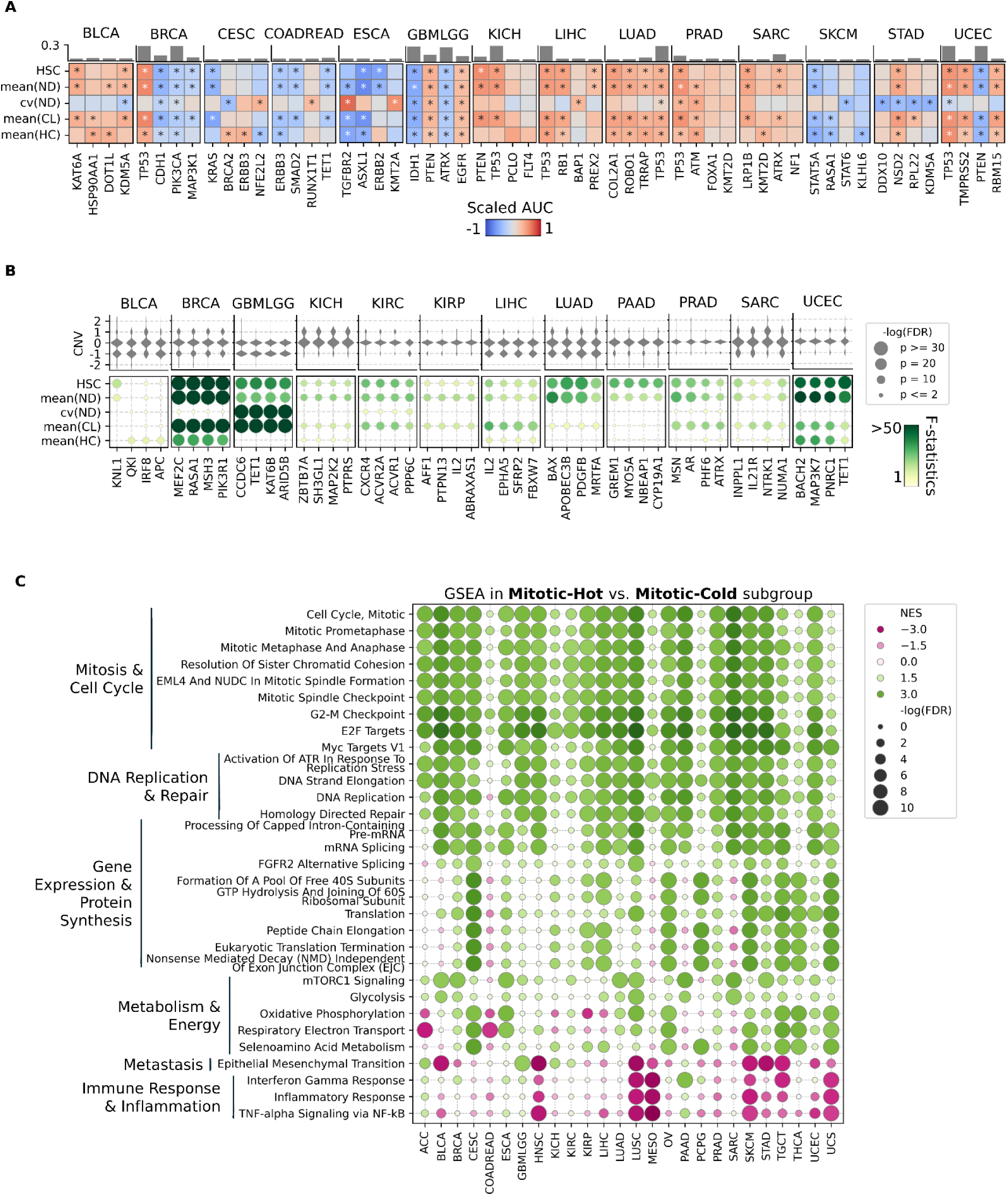
Mitotic topological features associate with the genomic alterations. (A) Association between MTFs and genes mutation status based on scaled-AUC (2*(AUC-0.5)). The bar plot on top of each gene column represent the percentage of population with that mutation within the related cohort. Asterisks indicate significant associations based on permutation test (q-value<0.05). (B) ANOVA analysis based on MTFs across different CNV states. The violin plot on top of each gene column represent the distribution of population of different CNV states of that gene across the related cohort. (C) Differentially expressed pathways between Mitotic-Hot and Mitotic-Cold groups based on GSEA.

Our analyses revealed that mutations in well-established tumor suppressor genes commonly align with expected changes in mitotic activity. For instance, *TP53* mutations are significantly and positively associated with increased mitotic density (mean(ND) and mean(CL)) across multiple cancers (BLCA, BRCA, COADREAD, ESCA, GBMLGG, KICH, LIHC, LUAD, LUSC, OV, PRAD, STAD, UCEC), reflecting the known loss of cell cycle control and enhanced proliferation when *TP53* function is compromised^23^. Similarly, *PTEN* mutations display positive associations with higher mitotic density in GBMLGG, KICH, and STAD which is consistent with unchecked PI3K/AKT pathway signaling in the absence of PTEN’s suppressive function^24^. In contrast, *PTEN* mutations in UCEC show a negative association with MTFs, suggesting *PTEN* mutations can be associated with less proliferative endometrial cancer^25^. Furthermore, *IDH1* mutations in GBMLGG, commonly linked to more indolent tumor behavior, also exhibit a negative association with mitotic density, aligning with the lower proliferative activity characteristic of *IDH1*-mutated gliomas^22^.

Beyond density, certain mutations correlate with mitotic heterogeneity (cv(ND)), including *BRCA2* (CESC/-, COADREAD/+), *KMT2A* (ESCA/+, UCEC/+), *BAP1* (KIRC/+, LIHC/+), *POLQ* (LUSC/-), and *PML* (SKCM/-). While these findings highlight gene-specific mutation effects on how mitoses distribute spatially, their mechanistic underpinnings are less established. Finally, the proximity of MAN (mean(HC)) shows gene- and cancer-type-dependent associations. For *HSP90AA1* (BLCA/+), *ERBB3* (CESC/+, COADREAD/-, UCEC/+), *KRAS* (CHOL/-, UCEC/-), *FAT1* (CESC/-, KIRP/-, LUSC/-), *NCOR2* (OV/+), and *NRAS* (THCA/+), patterns vary across tumor types, indicating that certain mutations are associated with either tight clustering or diffuse dispersal of mitotic cells within TME. These differences underscore the context-dependent nature of tumor evolution and the spatial dynamics of mitosis in diverse molecular backgrounds.

#### MTFs show associations with CpG sites linked to mitotic activity

Our Spearman correlation analyses (Table S4) demonstrate significant associations between MTFs and the methylation of different CpG sites across different cancers. In BRCA, MTFs show a strong negative correlation with methylation of CpG sites that are related to mitotic activity, notably TNFRSF10A (Spearman’s ρ: HSC -0.47, mean(ND) -0.48, mean(CL) -0.48). TNFRSF10A encodes TRAIL-R1, a key apoptosis regulator; its hypermethylation leads to epigenetic silencing, reduced apoptosis, and accumulation of mitotic cells^26^. This highlights that MTFs reflect epigenetic alterations affecting mitotic activity and apoptosis, and supports the therapeutic relevance of targeting TRAIL receptors. In MESO, we observed high negative correlation between MTFs and HTRA1 (mean(ND)=-0.54), a prognostic factor known to be positively associated with favorable survival outcomes^27^.

#### ANOVA analysis reveals association of MTFs with gene copy number variation

We assessed MTFs against gene-level copy number variation (CNV) states (deep deletion, shallow deletion, diploid, gain, amplification) using ANOVA (Figure 3B and Table S5), followed by Tukey’s post-hoc tests (Figure S4A).

Our analyses reveal robust associations between CNVs and mitotic activity across multiple cancers, with particularly strong effects in BRCA, GBMLGG, LUAD, PRAD, and UCEC. In BRCA, higher mitotic density, clustering, and proximity occur in deletion states of genes like *MEF2C*, *RASA1*, *MSH3*, and *PIK3R1*, suggesting loss of tumor-suppressive or genomic-stabilizing functions^28^. In GBMLGG, deletions in *KAT6B*, *TET1*, and *CCDC6* similarly correlate with increased mitotic density and clustering, pointing towards disrupted epigenetic regulation^29^. In LUAD, amplifications in *PDGFB* and *APOBEC3B* align with enhanced proliferation^30^. PRAD shows higher mitotic activity linked to alterations in *AR*, *PHF6*, *ATRX*, and *MSN*, highlighting the role of androgen signaling and chromatin remodeling in driving proliferation^31^. Finally, in UCEC, shallow deletions affecting *MAP3K7*, *BACH2*, and *PNRC1* also significantly correlate with increased mitotic activity, highlighting their tumor suppressive function^32,33^. Overall, most patterns align with established oncogenic or tumor-suppressive functions, some observed in a context never seen before.

#### Gene set enrichment analysis reveals pathways related to tumor proliferation

Gene set enrichment analysis (GSEA)^34^ on the differentially expressed genes (Figure S5) between Mitotic-Hot and Mitotic-Cold groups discovers differentially expressed pathways (Figure 3C and Table S6).

Pathways related to mitosis, cell cycle, and DNA replication are positively enriched across all cancers. This is due to their roles in driving rapid cell division, maintaining genomic stability under replication stress, and facilitating DNA repair, all of which are essential for sustaining the high mitotic activity and unchecked proliferation characteristic of aggressive tumor cells^35^.

In the Mitotic-Hot tumors, GSEA consistently highlighted mRNA processing, ribosome biogenesis, and translation machinery (Figure 3C), underscoring the heightened demand for protein synthesis in rapidly dividing cells. Notably, the “FGFR2 Alternative Splicing” pathway emerged as significantly enriched in CESC, LUSC, SKCM, STAD, TGCT, and UCS, pointing to a potential vulnerability in splicing-driven proliferation that extends prior reports of FGFR2 isoform switching^36^. Moreover, the “NMD Independent Of Exon Junction Complex (EJC)” pathway showed strong enrichment in Mitotic-Hot subgroups of CESC, KIRP, LIHC, OV, PCPG, SKCM, TGCT, THCA, and UCS, pointing to a potential role for NMD inhibition in restraining proliferation in these cancers^37^.

A proliferation-supporting pathway that has been enriched across all cancers is “mTORC1 signaling”, known to enhance anabolic processes and nutrient utilization^38^. The enrichment of the “respiratory electron transport” and “oxidative phosphorylation” pathways reflects the increased mitochondrial ATP production to meet tumor energy demands^39^ while the simultaneous upregulation of “selenoamino acid metabolism” suggests enhanced antioxidant defenses against Reactive Oxygen Species generated by intensified metabolic activity^40^.

Conversely, in cancers like ACC, LIHC, PAAD, and SARC, highly proliferative tumors downregulate the “oxidative phosphorylation” pathway and upregulate the “glycolysis” pathway – embracing the Warburg Effect^41^ – to efficiently produce energy and biosynthetic precursors through aerobic glycolysis while adapting to hypoxic conditions^39^. Activation of both glycolysis and oxidative phosphorylation in CESC, ESCA, LUSC, OV, UCEC, and UCS indicates generation of more ATP per glucose through combined processes supporting higher proliferation rates. This is inline with the findings of Hsu *et al.*^42^ that suggest both glycolysis and oxidative phosphorylation can coexist in tumor cells. This reliance on specific energy sources and metabolic processes can present new therapeutic avenues to disrupt tumor growth and proliferation^38–40^.

A negative enrichment of the “epithelial-mesenchymal transition (EMT)” pathway in Mitotic-Hot cases of BLCA, HNSC, LUSC, MESO, SKCM, STAD, TGCT, and UCEC is aligned with the "Go-or-Grow" hypothesis, which posits that cancer cells alternate between proliferative and migratory states^43^. On the contrary, positive enrichment of the EMT pathway in Mitotic-Hot cases across ACC, GBMLGG, and KICH, suggests that these tumors maintain their ability to proliferate while also exhibiting high invasiveness as reported before^44,45^.

#### PACMAN identifies druggable genes related to mitosis

The PACMAN genes were input into the Drug-Gene Interaction Database (DGIdb)^46^, which aggregates information on gene products, drugs, and drug-gene interactions to provide therapeutic insights (Figure S4B). Notably, TOP2A emerged as a key therapeutic target (whose gene has high correlation of r=0.64 with mean(ND) and mean(CL)). This protein, essential for DNA replication and chromosome segregation, is frequently overexpressed in aggressive cancers and is targeted by established drugs such as Etoposide, Paclitaxel, and Doxorubicin^47^. Combination strategies, such as dual inhibition of TOP2A and EZH2 (*EZH2* also correlated with MTFs, r=0.62), have shown enhanced efficacy in preclinical studies^48^.

Our analysis also highlights AURKA, AURKB, and PLK1, key mitotic regulators with known inhibitors capable of inducing mitotic arrest and apoptosis^49^. Additionally, *NEK2*, strongly correlated with mean(ND) (r = 0.66), represents another promising mitosis-specific target with therapeutic potential^50^.

### Prognostic significance of mitotic features

We performed pan-cancer survival analyses using recommendations and four endpoints defined by Liu *et al.*^11^ to assess the prognostic value of mitotic features: Progression-Free Interval (PFI), Disease-Specific Survival (DSS), Overall Survival (OS), and Disease-Free Interval (DFI).

Figure 4A presents heatmaps of C-indices across cancer types for DSS endpoints. Overall, mitotic features demonstrated heterogeneous prognostic performance across cancer types and endpoints. Several cancers (such as ACC, CHOL, and KIRC) showed significant C-indices for features like mean(CL), cv(ND), and mean(ND), with AMAH achieving 0.82 in KIRP. Atypical mitotic features (AMFs), including AMAH and AFW, consistently performed well in cancers such as KIRC, LIHC, LUAD, PAAD, and UCEC, highlighting their robust prognostic potential.

**Figure 4:**
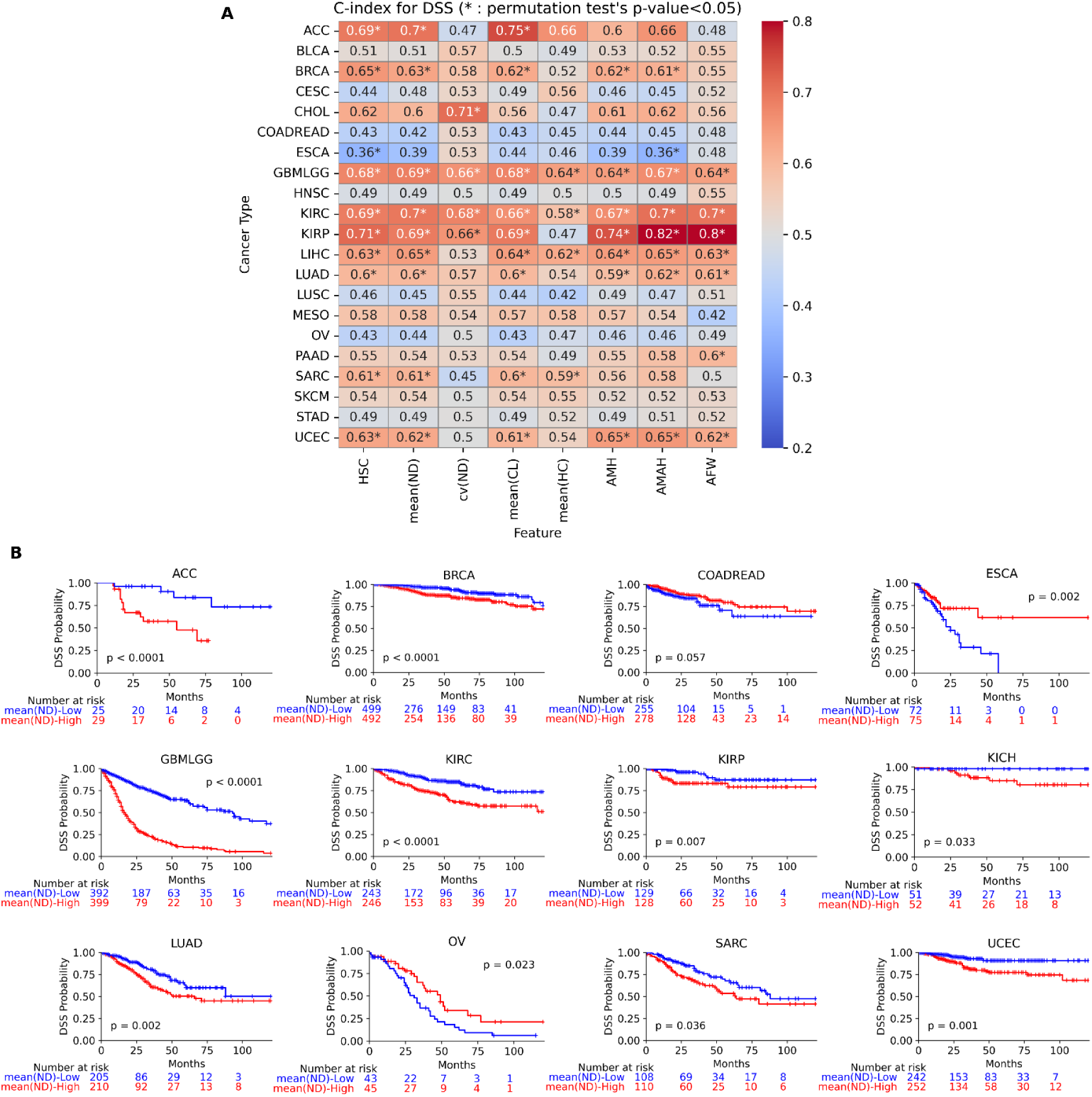
Mitotic topological features are prognostic biomarkers. (A) C-indices of MTFs and AMFs for DSS ranking. (B) Cross-validated KM curves^51^ for DSS across mean(ND)-Low and mean(ND)-High patients, where p-values show statistical significance through permutation tests.

While high mitotic activity typically indicates worse prognosis^8,52–54^, ESCA showed an inverse trend, where features like HSC and AMAH yielded C-indices below 0.5, suggesting aggressive behavior may instead be linked to reduced proliferation^55,56^. Similar patterns were noted, though less significant, in CESC, LUSC, OV, and COADREAD.

Comparable results emerged for other endpoints (Figure S6A–C). For PFI, significant C-indices were observed in 16 cancers, including BRCA, ESCA, GBMLGG, KIRC, LUAD, and SARC. Notably, in COADREAD, only cv(ND), which is a topological heterogeneity metric, achieved significant C-index, indicating its importance when density-based features fall short. The inverse proliferation–survival trend also appeared for PFI in ESCA and LUSC, with mean(HC) reaching a significant C-index of 0.42 in LUSC.

In the OS analysis (Figure S6B), significant prognostic associations were observed for many cancers, with mean(CL) reaching C-index of 0.75 in ACC and mean(HC) achieving 0.43 for LUSC. However, for DFI (Figure S6C), significance of mitotic features was confined to KIRP and PAAD with AMAH and LIHC with mean(CL), further underscoring the prognostic relevance of atypical mitosis.

To further validate feature performance, we conducted five-fold cross-validation of Kaplan-Meier (KM) curves^51^ using *mean(ND)* for patient stratification. Significant stratification was achieved for DSS in 11 cancers (Figure 4B, including ACC, BRCA, GBMLGG, KIRC, LUAD, SARC, and UCEC). Similar results were observed for PFI, OS and DFI in a subset of these (Figure S7A-C). In ESCA and OV, mean(ND)-Low groups showed significantly poorer survival, reiterating inverse trends noted in the C-index analysis.

Although *mean(ND)* was not predictive in several cancers (e.g., BLCA, LUSC, SKCM), other features, such as AFW, demonstrated potential prognostic value in these contexts (Figure 5C). This supports further exploration of both MTFs and AMFs across tumor types.

**Figure 5:**
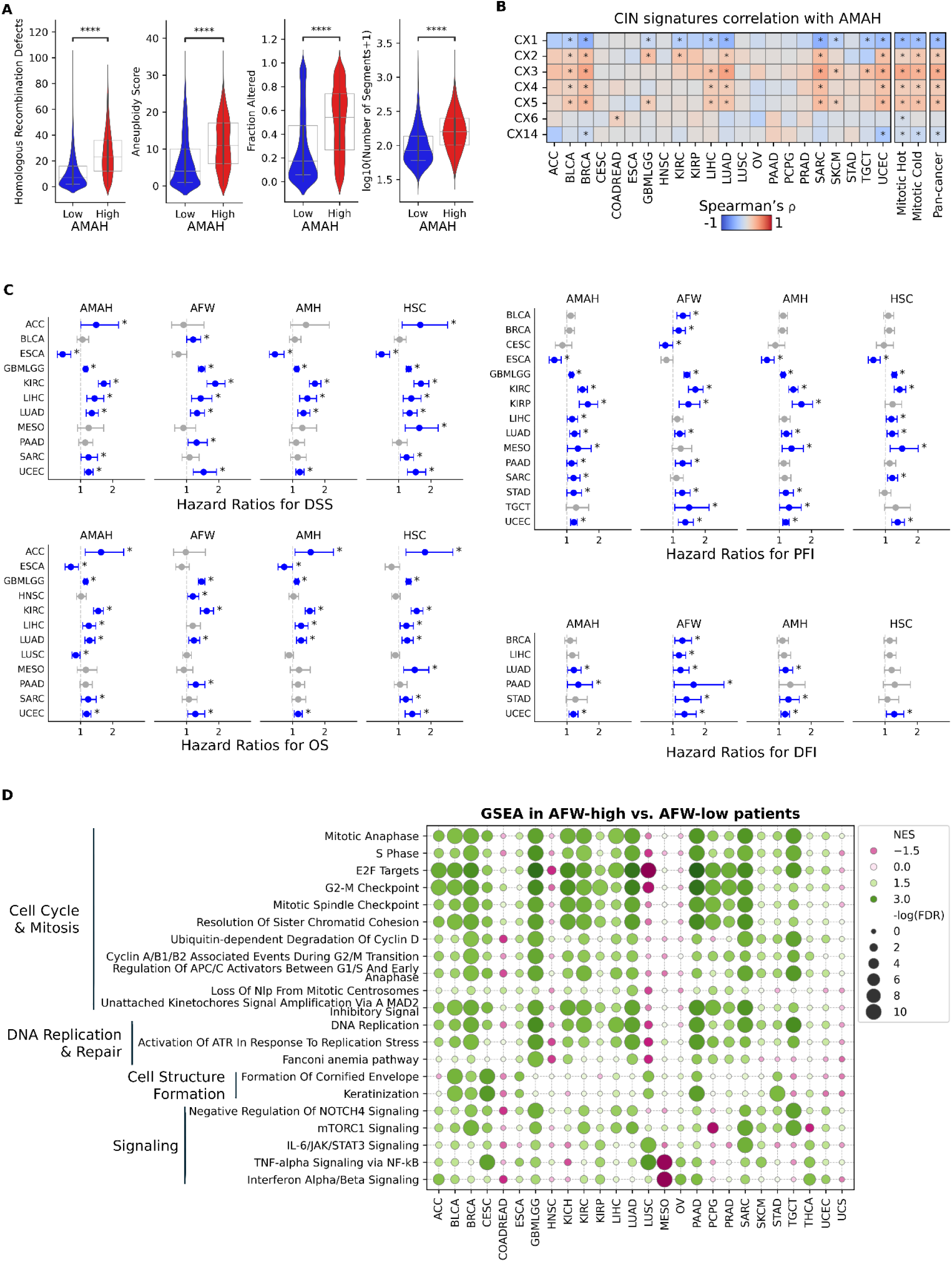
Measuring mitotic errors through atypical mitotic activity. (A) Violin plots comparing tumor characteristics between AMAH-High and AMAH-Low groups on pan-cancer level. Asterisks denote significance levels of the two-way Mann-Whitney U test (****:p≤0.0001). (B) Heatmap of Pearson’s correlation (r) between AMAH and CIN signatures^58^. Asterisks indicate significant correlations (q-value<0.05). (C) Hazard ratios of different AMFs and HSC, taken from univariate survival analysis across different cancer and event types. For each cancer, Asterisks denote significant stratification of patients (log-rank test p-value<0.05) based on the median value of each feature. (D) Differentially expressed pathways between AFW-High group vs. AFW-Low group based on GSEA.

### Measuring mitotic errors through atypical mitotic activity

#### The higher the mitotic activity, the greater the number of mitotic errors

We investigate the distribution of AMAH across different cancers, genders, races, and mitotic subgroups (Figure S8A) where we found a similar trend to average mitotic activity across cancers (Figure 2C). Furthermore, in Figure S8B, we found that the higher the mitotic activity (HSC), the higher the atypical mitotic activity (AMAH) with Pearson’s *r*=0.84. One explanation is that high mitotic activity in tumors overwhelms quality control mechanisms, leads to the accumulation of genetic mutations, increases stress on mitotic processes, involves defective apoptosis mechanisms, and results in chromosomal instability (CIN), all contributing to a higher incidence of atypical mitoses^1,2^.

#### AMFs are associated with genomic instability

CIN is often characterized by a high rate of chromosome missegregation during cell division among other reasons^57,58^. CIN and mitotic errors have a bidirectional relationship, where each can exacerbate the other, and this instability often results in increased tumorigenesis and aneuploidy in cancer^1,59–61^. This effect is illustrated in Figure 5A, where the pan-cancer distributions of Homologous Recombination Defects (HRD), aneuploidy, DNA fraction alteration, and the number of segments in the AMAH-High group are all significantly higher than in the AMAH-Low group (High and Low groups are defined based on the median AMAH cutoff).

Leveraging the seminal work of Drews *et al*.^58^, we can measure the association of our AMFs with different CIN signatures. Our findings (Figure 5B) suggest that CX1 and CX14 signatures have significant inverse correlation with AMAH at pan-cancer level (Spearman’s *ρ* values of -0.4 and -0.11, respectively) whereas CX2-CX5 signatures correlate positively significant with AMAH (Spearman’s *ρ* values of 0.2, 0.4, 0.2, and 0.27, respectively). The observed associations between AMAH and CIN signatures reflect distinct mechanisms of CIN influencing tumor cell mitosis. CX1 and CX14 measure chromosomal missegregation events driven by telomere dysfunction and reduced telomerase activity^58^. These telomere-related defects can trigger cellular senescence or apoptosis due to DNA damage responses from critically short telomeres^62^, limiting mitotic activity and resulting in fewer observable atypical mitoses, explaining their negative association with AMAH. In contrast, CX2, CX3, and CX5 capture replication stress and HRD, leading to catastrophic genomic instability characterized by tandem duplications and loss of heterozygosity^58^. Replication stress causes unresolved DNA replication intermediates and under-replicated regions, resulting in visible mitotic errors like bridge formation and multipolar spindles^63,64^, thereby increasing detectable atypical mitoses and justifying their strong positive association with AMAH. Similarly, CX4 reflects whole-genome duplication^58^, which often causes centrosome amplification and multipolar mitoses (tetraploid cells)^65^, contributing to detectable high AMAH activity.

#### AMFs are independent prognostic biomarkers

From univariate survival analysis of AMAH, AFW, AMH, and HSC features (Figure 5C), we note that wherever standard HSC was not significant in stratification, one of AFW, AMAH, or AMH gave significant risk stratification (e.g., in BLCA, BRCA, CESC, KIRP, PAAD, STAD, and TGCT for PFI). Similar trend was discussed before in Figures 4A and S6 for C-index in survival ranking. It is observed that AFW may be preferrable over other measures for PFI and DFI events. In particular, AFW gives significant stratification of BRCA, LIHC, LUAD, PAAD, STAD, and UCEC for DFI, whereas HSC can only stratify patients in UCEC (Figure 5C). This highlights the potential importance of quantifying atypical mitoses instead of general mitotic activity in clinical settings, where DFI and PFI are key clinical endpoints in guiding decisions for treatments. From Figure 5C, higher atypical mitotic activity is associated with worse prognosis in almost all cancers, except for CESC and ESCA.

Multivariate survival analyses were also conducted to evaluate the independent prognostic value of AFW, AMAH, and AMH (Tables S7-S10). We observe that AFW is more frequently an independent predictor of survival among the three AMFs (Table S7) and can add prognostic value beyond standard HSC in many cancers (Table S8), including BLCA, CESC, COADREAD, GBMLGG, KIRC, KIRP, LUAD, PAAD, STAD, and UCEC.

To further illustrate the prognostic values of AMFs regardless of mitotic count, we performed patient stratification based on median AFW value in Mitotic-Hot and Mitotic-Cold subgroups (Figure S8C). In Mitotic-Hot group, patients with lower AFW showed significantly better PFI probability in KIRC, KIRP, and UCEC and better DSS probability in LIHC. Furthermore, PFI probabilities for KIRC, PAAD, and TGCT as well as DFI probability for LUAD are significantly better in AFW-Low patients in the Mitotic-Cold subgroup. These findings suggest that even in patients with low proliferative activity, the presence of atypical mitoses is indicative of worse prognosis, underscoring the need for closer monitoring and more aggressive treatment strategies.

#### GSEA reveals genomic pathways related to mitotic errors

GSEA is performed between AFW-High and AFW-Low patients to identify pathways associated with higher rates of mitotic errors (Figure 5D and Table S11). The upregulation of “Cell Cycle & Mitosis” pathways (especially the first six pathways in Figure 5D) across most cancers suggests heightened cell proliferation and activation of cell cycle checkpoints in response to higher mitotic errors and genomic instability^1,2,61,63^. More specifically, positive enrichment of pathways related to the degradation of Cyclin D or presence of Cyclin B, regulation of APC/C activators, unattached kinetochores signaling, and loss of Nlp from mitotic centrosomes all align with the potential reasons explained in the literature for increased mitotic errors^1,2,66–68^.

There are well established links between DNA replication stress and mitotic errors such as chromosome missegregation^1,69,70^, the evidence of which is also presented in Figure 5D where DNA replication and activation ATR in response to replication stress is significantly upregulated in AFW-High patients in most cancers (specifically, BRCA, GBMLGG, KIRC, LUAD, PAAD, SARC, and TGCT). Additionally, the “Fanconi Anemia” pathway, which has been linked to weak spindle assembly checkpoint and mitotic errors^71^, was also found to be upregulated in AFW-High patients in GBMLGG, KICH, LUAD, and PAAD, indicating increased DNA damage and activation of repair mechanisms^71^. These dependencies of cancer on DNA repair pathways offer potential therapeutic targets^72,73^.

In our study, we observed that pathways involved in cornified envelope formation and keratinization were positively enriched in patients with higher rates of mitotic errors in BLCA, BRCA, CESC, ESCA, LUSC, PAAD, and STAD. The upregulation of these pathways in cancer cells may lead to cytoskeletal alterations and interfere with mitotic spindle formation, increasing the likelihood of mitotic errors^74^. Additionally, the aberrant activation of these cytoskeletal alterations can disrupt normal cell cycle regulation and checkpoints, further contributing to genomic instability^75^.

On investigating other important signaling pathways (Figure 5D), we observe enrichment of the “negative regulation of NOTCH4 signaling pathway” in AFW-High group in most cancers. This pathway reduces NOTCH4 activity, which can modulate microtubule stability^76^. The altered microtubule dynamics result in mitotic defects such as chromosome misalignment and segregation errors^1,76^. The positive enrichment of the “TNF-α/NF-κB” and “IL-6/JAK/STAT3” signaling pathways in the AFW-High group of most cancers (especially CESC, LUSC, and SARC) stems from their cooperative role in driving cell proliferation and survival, as their pro-inflammatory effects can exacerbate CIN by creating a tumor-supportive environment that promotes unchecked cell division and shields cells from apoptosis^1,77,78^. Similarly, “interferon α/β” signaling contributes to a pro-inflammatory tumor microenvironment that promotes tumor progression, CIN, and mitotic errors^79–81^. However, in MESO, we observed a downregulation of these pathways in AFW-High cases. This divergence highlights the unique biology of mesothelioma and underscores the importance of considering cancer-specific mechanisms when developing therapeutic strategies.

### Mitotic activity and the immune landscape of tumors

#### MTFs are associated with immune phenotypes of tumor

Spearman correlation analyses were performed between mean(ND) as a measure of mitotic activity and different immune cell expressions^14^ (Figure 6A), revealing significant correlations between mitotic activity and the expression levels of various T-helper (Th) cell subsets, alongside enrichment patterns (Figure 3C) that align with these findings. Specifically, Th1 cells exhibited a negative correlation with mitotic activity (*ρ*=-0.3; q-value<0.0001 on pan-cancer level), suggesting that higher Th1 cell presence is associated with reduced tumor proliferation. This aligns with the anti-tumor role of Th1 cells, which produce interferon-gamma (IFN-γ) to promote cytotoxic immune responses^82^. GSEA results (Figure 3C) with negative enrichment of the “Interferon Gamma Response” pathway in Mitotic-Hot group across most cancers further confirms this. Moreover, downregulation of “TNF-alpha signaling via the NF-κB” pathway in the Mitotic-Hot group (Figure 3C) can be linked to reduced Th1 cell expression with higher mitotic activity, because NF-κB has a critical role in Th1 differentiation and function^83^. Similar to Th1, Th17 cells also showed a negative correlation with mitotic activity (*ρ*=-0.27; q-value<0.0001 on pan-cancer level), indicating their potential involvement in anti-tumor immunity through inflammatory responses and recruitment of effector cells such as activated natural killer cells^84^, which also have a lower expression in cases with higher mitotic activity (*ρ*=-0.09; q-value<0.0001 on pan-cancer level). This functionality of Th17 cells can also be due to their plasticity which allows them to polarize towards a Th1-like phenotype in vivo^82,85^. In contrast, Th2 cells demonstrated a strong positive correlation with mitotic activity; the highest among all immune expressions on pan-cancer level with *ρ*=0.53 and significantly positive across almost all cancers, except for THYM. This implies that elevated Th2 responses are associated with increased tumor proliferation. This supports the role of Th2 cells in creating a tumor-promoting environment via immunosuppressive cytokines (like IL-4 and IL-13) and inhibiting Th1 responses^82,86^. Significantly higher distribution of Th1:Th2 cells ratio in Mitotic-Cold group (Figure 6B) further confirms this trend. Additionally, regulatory T cells (Tregs) showed a positive correlation with mitotic activity (*ρ*=0.14; q-value<0.0001 on pan-cancer level), which may be due to the interplay of Treg and Th cells where increased Treg presence can suppress anti-tumor immune responses (Th1 and Th17) but not Th2^85,87^ and therefore contributing to tumor proliferation. Moreover, we found the “Lymphocyte Infiltration Signature” to be significantly higher in the Mitotic-Cold vs. Mitotic-Hot group on pan-cancer level (Figure 6B).

**Figure 6:**
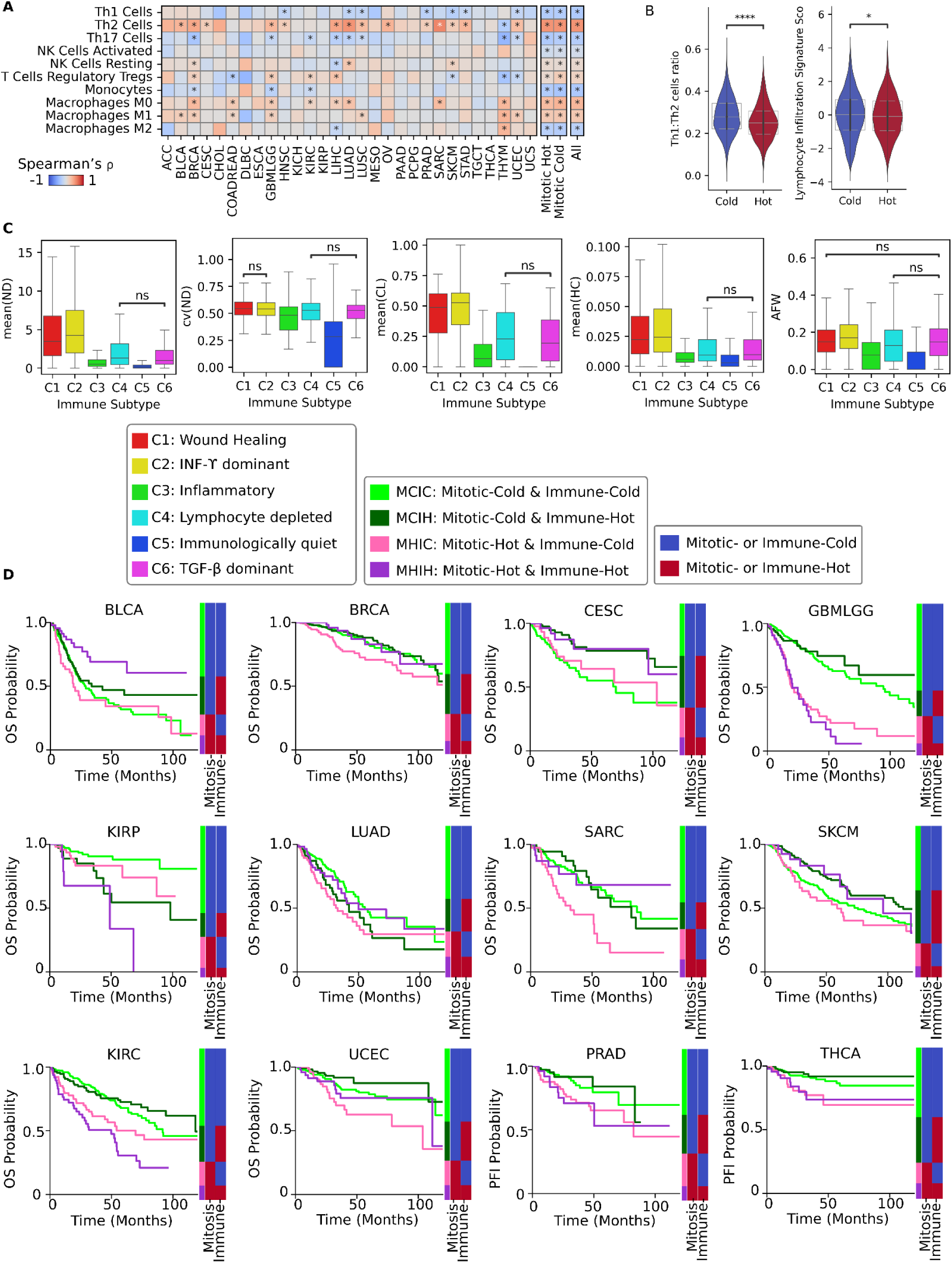
Mitotic activity and the immune landscape of tumor. (A) Heatmap of Spearman’s correlation (ρ) between immune expressions and mean(ND), Asterisks indicate q-values≤0.05. (B) Distribution of tumor’s immune properties over Mitotic-Cold and Mitotic-Hot groups (Mann-Whitney statical test, *:p<0.05, ****:p<0.0001). (C) Distribution of MTFs and AFW over different immune subtypes (except for the pairs annotated as “ns”, all other pairs are significantly different based on Mann-Whitney statical test, p<0.05). (D) Patient stratification based on two variables: mitotic temperature and immune (CD8 T cells expression) temperature. Significant stratifications are explained in the main text. Here, stacked bars represent the population proportion for different subgroups in each cancer type.

However, some findings did not align with the established roles of the immune cells. Notably, macrophage M1 cells showed a positive correlation with mitotic activity (*ρ*=0.22), which is not in line with their recognized function in anti-tumor immunity through pro-inflammatory actions^88^. Conversely, macrophage M2 cells displayed a negative correlation (*ρ*=-0.22) with mitotic activity across the pan-cancer cohort, despite generally being associated with tumor promotion due to their immunosuppressive and tissue remodeling activities^88^. Nevertheless, we found that in MESO and THYM, macrophage M2 cells had a significant positive correlation with mitotic activity. Moreover, our findings reveal a negative correlation between monocyte expression and mitotic activity (*ρ*=-0.3), which may appear to contrast with the evidences of monocytes generally promoting tumor growth^89^.

#### Different immune subtypes show different levels of mitotic activity

Box plots in Figure 6C show that both mitotic activity and atypical mitotic activity (AFW) vary significantly across the different immune subtypes proposed by Thorsson *et al.*^14^. Mitotic density (mean(ND)), mitotic clustering (mean(CL)), mitotic closeness (mean(HC)), and mitotic errors (AFW) were highest in the C2 (IFN-γ dominant) and C1 (Wound Healing) immune subtypes, which also have relatively higher proportion of Mitotic-Hot cases (Figure 1D). In C2, the elevated mitotic activity and higher median AFW may be due to high proliferation rates and genomic instability, possibly resulting from mechanisms that allow tumors to evade immune destruction or from the immune response inducing selection of more proliferative tumor clones. Similarly, in C1, the high Th2 cell bias and increased wound healing processes, such as elevated angiogenic gene expression, may have led to tumor growth, increased mitotic activity, and elevated mitotic errors^21^. Subtypes C4 (Lymphocyte Depleted) and C6 (TGF-β dominant) showed intermediate levels of both mitotic activity and atypical mitotic activity (albeit AFW levels in C4 and C6 are more close to C1 and C2). The moderate mitotic activity in these subtypes could be related to their immunosuppressive environments (C4 with suppressed Th1 activity and C6 with elevated TGF-β levels) that allow some tumor proliferation and accumulation of mitotic errors but not at the highest levels observed in C1 and C2. The lowest mitotic activity and mitotic errors were observed in C3 (Inflammatory) and C5 (Immunologically Quiet) subtypes. In C3, elevated Th1 and Th17 responses may enhance anti-tumor immunity and suppress tumor proliferation. C5 was notably mitotically quiet, which aligns with its characteristics of low proliferation rates, low genomic instability, and association with less aggressive tumors, like lower-grade gliomas, suggesting that inherent tumor properties contribute to its reduced proliferation and lower rates of mitotic errors. Additionally, mitotic distribution heterogeneity (cv(ND)) was similar across all subtypes except for C5, which had significantly lower heterogeneity than the rest, further highlighting its distinct mitotically quiet behavior.

#### Combined effect of mitotic activity and CD8 T cells expression on survival

We analyzed the combined impact of mitotic activity and immune activity (measured by CD8+ Cytotoxic T Lymphocytes or CTLs) on patient survival across various cancers (Figure 6D) to better understand their interplay and identify potential therapeutic implications. To this end, patients within each cancer type were categorized into four groups based on their mitotic activity and immune activity temperatures:

- MCIC: Mitotic-Cold & Immune-Cold
- MCIH: Mitotic-Cold & Immune-Hot
- MHIC: Mitotic-Hot & Immune-Cold
- MHIH: Mitotic-Hot & Immune-Hot

KM survival curves were plotted for each group, and pairwise log-rank tests were performed to assess statistical significance.

In some cancers, such as BLCA, BRCA, CESC, SARC, SKCM, and UCEC, higher CTLs expression improved OS despite high mitotic activity. In BLCA, the MHIH subgroup showed significantly better OS compared to the other subgroups, with p-values of 0.002 against MCIC, 0.035 against MCIH, and 0.0009 against MHIC. This suggests that a robust CTLs response can improve survival even in highly proliferative tumors. In BRCA, the MHIC subgroup had significantly worse OS compared to the other groups, with p-values of 0.007, 0.010, and 0.039 against MCIC, MCIH, and MHIH, respectively. Similarly, in SARC, the MHIC subgroup showed significantly worse OS, with p-values of 0.0003, 0.003, and 0.025 against MCIC, MCIH, and MHIH, respectively. For UCEC, the most significant stratification was between MCIH and MHIC groups (p-value=0.0002), with MHIC showing worse prognosis. These findings indicate that high mitotic activity coupled with low CTLs expression leads to poorer outcomes, while high immune activity can counteract the negative impact of high proliferation. In CESC and SKCM, Immune-Hot subgroups (MCIH and MHIH) demonstrated better survival than Immune-Cold subgroups (MCIC and MHIC), regardless of mitotic activity. This suggests that CTLs-mediated immune responses play a crucial role in controlling tumor progression in these cancers. These cancers, characterized by inherent immunogenic features^90^, may particularly benefit from immunotherapy strategies, such as immune checkpoint inhibitors, that enhance CTLs function^91^.

Conversely, in GBMLGG, KIRC, PRAD, and THCA cancers, where high mitotic activity worsened prognosis regardless of CTLs expression, pointing to the possibility that these cancers are less immunogenic. In GBMLGG, Mitotic-Cold subgroups (MCIC and MCIH) had significantly better OS than Mitotic-Hot subgroups, with p-values<0.00001 for any pairs comparing Mitotic-Cold to Mitotic-Hot, regardless of CTLs expression. KIRC showed a similar pattern for OS, while PRAD and THCA exhibited a similar trend for PFI. This suggests that high proliferation is the key factor affecting prognosis in these cancers, and targeting mitotic activity^49,50,92,93^ may be therapeutically beneficial.

Interestingly, in KIRP and LUAD cancers, high CTLs expression was associated with poor prognosis. In KIRP, within the Mitotic-Cold subgroups, MCIH showed significantly worse survival compared to MCIC (p-value=0.001). Similarly, within the Mitotic-Hot subgroups, MHIH had worse prognosis than MHIC (p-value=0.012). This suggests that in KIRP, higher CTLs expression may be associated with dysfunctional or pro-tumor immune responses. In such cancers, therapeutic strategies might focus on enhancing the functionality and survival of CTLs within the immune microenvironment to restore effective anti-tumor immunity^94^. Furthermore, in LUAD, MCIC showed significantly better OS compared to MCIH (p-value=0.041) and MHIC (p-value=0.005). This indicates that increased CTLs expression or higher mitotic activity separately worsen prognosis, but when both are highly activated (MHIH), survival was not significantly worse than MCIC. This complex interplay suggests that the presence of CTLs alone may not be sufficient, possibly due to immune suppression or exhaustion within the tumor microenvironment. Further investigation is needed to understand the mechanisms behind this observation and to develop appropriate therapeutic strategies.

## DISCUSSION

This study underscores the critical role of mitosis as both a driver and a marker of tumor aggressiveness. By leveraging the MAN and SNA, we effectively capture topological features of mitotic activity that traditional approaches often overlook. Furthermore, for the first time, we comprehensively investigated the role of atypical mitosis (a surrogate for mitotic errors) in patient prognosis across many cancers. Our approach relies on efficient DL-based mitosis detection algorithms that have shown pathologist-level accuracy^95^, therefore, enabling cost-effective mitosis-related biomarker extraction from H&E slides with implications for risk stratification and precision oncology.

By investigating the interplay between mitotic activity, genomic alterations, and immune characteristics, this study shows that mitotic activity is closely linked to several hallmarks of cancer^96^. Increased mitotic activity reflects the ability of cancer cells to sustain proliferative signaling such as E2F and MYC targets (Figure 3C). The density of atypical mitoses significantly correlates with CIN^58^ (Figure 5A-B), enabling cancer evolution by promoting mutations and chromosomal abnormalities^2^. The relationship between high mitotic density, *TP53*, and *PTEN* mutations also outlines the association of growth suppressor evasion with tumor proliferation (Fig 3A and Table S3). Moreover, high mitotic activity is closely related to metabolic reprogramming in tumors, with context-dependent reliance on glycolysis and oxidative phosphorylation to sustain rapid cell division (Figure 3C). And finally, changes in the patterns of immune cell infiltration, including decreased Th1 responses and elevated Th2 activity in dividing tumors, point to a relation with immune evasion and tumor-promoting inflammation mechanisms (Figure 6A-B). These findings collectively point to the notion of mitotic activity as a pivotal nexus at which multiple hallmarks of cancer converge.

### Integration with established knowledge and emerging pan-cancer insights

Our findings, derived from both exploratory and confirmatory analyses, demonstrate that MTFs provide robust insights into tumor aggressiveness across diverse cancers.

A notable contribution is the identification of the PACMAN gene set (Table S2), which extends the existing gene set for mitotic activity (MNAI^17,18^) with novel candidates, providing a comprehensive resource for understanding mitotic regulation. Key PACMAN genes, such as *TOP2A*, *EZH2*, *AURKA*, and *PLK1* are known as druggable targets^47–49^.

Furthermore, we found significant differences in mitotic activity across cancers (Figures 2C and S2). For example, mitotic activity is notably higher in squamous cell carcinomas (e.g., CESC and LUSC), suggesting tissue-specific mitotic dynamics influenced by genetic and microenvironmental factors. Additionally, ethnicity differences in mitotic activity within cancers such as BRCA and LIHC point to underlying biological differences warranting further study.

### Novel clinical implications, contradictory results, and future directions

Our findings present significant clinical implications by demonstrating the utility of MTFs in refining prognostic models across diverse cancers. In particular, features such as mitotic connectivity (mean(ND)) and heterogeneity (cv(ND)) were shown to outperform HSC for survival ranking in a few cancers (Figures 4A and S6). We found previously unreported evidence of prognostic value for mitotic quantification across several cancers, including ACC, ESCA, PRAD, and notably kidney cancers (KIRC, KIRP, KICH). These insights indicate the potential clinical utility of MTFs in precision oncology. However, the underlaying mechanisms that derive mitotic heterogeneity are still unknown and require further investigation.

A key novel contribution of this study lies in elucidating the independent prognostic role of atypical mitotic features. The association between high AMFs and poor prognosis in most cancers (more pronounced for DFI) underscores the importance of monitoring mitotic errors as markers of aggressive tumor behavior.

The pathway analyses in this study provide novel insights into the complex biological mechanisms underlying tumor proliferation and mitotic errors, revealing critical dependencies that can inform precision oncology. The analyses reveal mechanisms central to tumor aggressiveness, highlight tumors’ adaptive metabolic flexibility, and link mitotic errors to DNA repair activity and cytoskeletal disruptions, all of which suggest shared and context-specific therapeutic avenues.

Our survival analyses based on mitotic and immune activity combined revealed interesting therapeutic avenues for different cancers. We found that for some immunogenic cancers (e.g., BLCA and BRCA), higher CTLs responses can lead to better survival regardless of their mitotic activity, and therefore patients might benefit from immune checkpoint inhibitors that enhance T cell-mediated cytotoxicity. Conversely, targeting mitotic regulators could disrupt proliferation in less immunogenic cancers (e.g., KIRC and PRAD).

Our findings present some contradictions too. For instance, in CESC and ESCA, our results showed association between higher (atypical) mitotic activity and better outcomes. However, most studies suggest that higher mitotic activity is associated with worse prognosis in ESCA and CESC^97,98^, although there is evidence in cervical cancer that mitotic activity is not necessarily directly associated with tumor grading or aggressiveness due to hormonal influences^99^. Nevertheless, our results can be an evidence that a high rate of CIN and mitotic errors in UCEC and ESCA (Figure S8A) may lead to tumor suppression or cell death^2,57^. This is a property that can be strategically targeted for therapy, using drugs like Paclitaxel, to push cancer cells beyond their tolerance for CIN and causing them to die^2^.

Similarly, discrepancies in M1 and M2 macrophage cells dynamics in relation to mitotic activity were observed. Furthermore, the observed context-dependent enrichment of EMT pathway in highly mitotic tumors challenges conventional dichotomies of proliferation and invasion, suggesting a nuanced interplay that varies by cancer type. Such insights can be exploited to identify new drug targets based on existing knowledge. For example, it is shown that CDK1 promotes simultaneous EMT and proliferation in ACC, while the inhibitor “cucurbitacin E” suppresses both^44^. Therefore, the effectiveness of this inhibitor can also be examined in GBMLGG and KICH where EMT and proliferation show similar relationship.

Future research may focus on unraveling the molecular mechanisms linking mitotic activity, immune modulation, and tumor heterogeneity. Multi-centric validation of mitotic features across different cancers and ethnicities using large-scale datasets is crucial to robustly examine the proposed MTFs and AMFs for clinical use.

### Limitations of the Study

The methodology and features designed in this study are specifically focused on mitotic activity and do not address possible interactions of mitoses with other cell types within the TME. These interactions could yield additional insights and present opportunities for future studies to build upon our resources.

Although we employed robust validation techniques, including external datasets and randomized slide checks (Materials and Methods), variations in slide preparation and imaging systems may introduce inaccuracies that affect mitosis detection and the robustness of derived features. Future improvements in mitosis detection and tumor segmentation models will allow further refinement of our approach.

Moreover, our work relies on diagnostic slides of TCGA, where some cancers (ACC, CHOL, DLBC, MESO, UCS) are underrepresented. Therefore, more comprehensive datasets on such rare cancers are required to further validate our findings. Furthermore, due to the large scale of this study, our analyses were limited to original TCGA project cohorts, each comprising multiple cancer subtypes. Future investigations leveraging our resources could focus on subtype-specific analyses of mitotic behavior to gain more nuanced insights.

Despite these limitations, this study reaffirms the critical role of mitotic activity in cancer biology and offers valuable resources for further research. The extensive analyses presented here serve as a demonstration of the potential applications of our methodology and resources, which can aid further investigations into the interplay of mitotic activity and TME and their joint impact on patient outcomes and finding suitable therapeutic targets.

## Supporting information

Table S1: Excel file containing mitotic features extracted from 9,141 cases across 31 cancers sourced from TCGA.

Table S2: Excel file containing list of PACMAN genes and their correlations with MTFs

Table S3: Excel file containing full results of measuring association of gene mutations with MTFs.

Table S4: Excel file containing full results of measuring association of methylation CpG with MTFs.

Table S5: Excel file containing full results of ANOVA analysis of MTFs association with CNV.

Table S6: Excel file containing full results of GSEA between Mitotic-Hot and Mitotic-Cold subgroups.

Table S7: Excel file containing full results of multivariate survival analysis using atypical mitotic features across all cancer and event types.

Table S8: Excel file containing full results of multivariate survival analysis using AFW and HSC.

Table S9: Excel file containing full results of multivariate survival analysis using AMAH and HSC.

Table S10: Excel file containing full results of multivariate survival analysis using AMH and HSC.

Table S11: Excel file containing full results of GSEA between AFW-High and AFW-Low subgroups.

## Data Availability

All data produced in the present work are contained in the manuscript.

https://github.com/mostafajahanifar/pacman/tree/paper/

## Resource availability

### Materials availability

The datasets and resources generated in this study are publicly available. The TCGA mitosis dataset and the mitosis subtyping dataset for atypical mitosis detection are hosted on Zenodo at https://zenodo.org/records/14548480 and https://zenodo.org/records/15390543, respectively. The TCGA mitotic features table, PACMAN gene set, and results from associated genomic and survival analyses are provided in Supplemental Tables S1–S11 available at Zenodo: https://zenodo.org/records/14793678. An online interactive platform for visualizing mitotic activity networks is available at https://tiademos.tia.warwick.ac.uk/bokeh_app?demo=pacman.

### Data and code availability

The TCGA diagnostic WSIs used in this study are publicly available through the Genomic Data Commons (GDC) Data Portal at https://portal.gdc.cancer.gov/. The initial dataset used to train the (atypical) mitosis detection models^100^ as well as all the clinical, genomic, and immunological datasets used in this manuscript are all publicly accessible (https://zenodo.org/records/15603656).

All original code used for reproducing all analyses, figures, and tables in this study has been deposited at GitHub and is publicly available at https://github.com/mostafajahanifar/pacman. Any additional information required to reanalyze the data reported in this paper is available from the corresponding author upon reasonable request.

## Acknowledgments

Authors would like to thank Dr. Asmaa Ibrahim for kindly reviewing the image patches for mitosis subtyping dataset. MJ, SR, and NR collaborate with the BigPicture consortium (www.bigpicture.eu), which has received funding from the Innovative Medicines Initiative 2 Joint Undertaking under grant agreement No 945358. This Joint Undertaking receives support from the European Union’s Horizon 2020 research and innovation program and EFPIA (www.imi.europe.eu). The results in this paper are in whole or part based upon data generated by the TCGA Research Network (https://www.cancer.gov/tcga). CAB acknowledges the support from the Austrian Research Fund (FWF, project number: I 6555). MA acknowledges support by the Deutsche Forschungsgemeinschaft (DFG, German Research Foundation, project number: 520330054).

## Author contributions

M.J. conceptualized the methodology, designed the study and experiments, performed model evaluation and analyses, generated figures, and wrote the manuscript. M.D. provided genomic data, consulted on genomic analyses, and contributed to writing. N.Z. implemented the mitotic activity network and topological features and contributed to writing. A.S., B.S.C., and C.A.B. supported data labeling, and clinical interpretation. N.W. assisted with survival analysis design. M.W. implemented the online visualization platform. M.A. provided datasets. S.E.A.R. designed immune landscape analyses. F.M. reviewed and revised analyses. N.R. contributed to conceptualization and supervised the project. A.S., B.S.C., C.A.B., N.W., M.A., S.E.A.R., F.M., and N.R. edited the manuscript.

## Materials and Methods

### Data inclusion protocols

The Cancer Genome Atlas (TCGA) project^12^ served as the source of data for this study. Figure 1A summarize the TCGA studies and the number of cases included in this chapter, in which official abbreviations of TCGA studies are utilized (https://gdc.cancer.gov/resources-tcga-users/tcga-code-tables/tcga-study-abbreviations). Diagnostic slides were downloaded directly from the Genomic Data Commons data portal (https://portal.gdc.cancer.gov). Cases without diagnostic slides were excluded. Subsequently, slides were reviewed in an automated fashion to exclude cases without complete diagnostic slides (WSIs that missing appropriate metadata) or clinical information^11^. For cases with multiple WSIs, the slide with the highest number of mitoses was selected to represent the case. TCGA studies related to non-solid tumors (e.g., LAML and LCML) were excluded, as well as Uveal Melanoma (UVM) due to the extensive DAB staining observed in its slides, which could compromise mitosis detection accuracy. Finally, genomic, transcriptomic, and epigenetic data for these samples were collected from cBioPortal^13^.

Note that the COAD and READ projects were combined into COADREAD, and GBM and LGG were merged into GBMLGG to ensure comprehensive analysis of colorectal and brain tumors, respectively. In total, 9,141 tumors across 31 cancer types were included in our pan-cancer analysis.

### Mitosis detection algorithm

We utilized a two-stage framework called “Mitosis Detection, Fast and Slow” (MDFS) ^15^. The first stage, termed "Detecting Fast", uses a segmentation convolutional neural network to quickly and sensitively identify mitotic candidates at reduced resolution. The second stage, "Detecting Slow", refines these candidates using a deep classifier network operating on high-resolution patches to differentiate mitoses from mimickers. To address domain shift issues common in histology images (such as stain and resolution variation), the methodology integrates domain generalization techniques, including stain normalization and augmentation. Our approach demonstrated state-of-the-art accuracy and generalizability across various domains, winning two major challenges on mitosis detection MIDOG21^101^ and MIDOG22^95^.

#### Validation of mitosis detection

During the training, the algorithm has seen images from human breast carcinoma, canine lung cancer, canine lymphoma, canine cutaneous mast cell tumor, and human neuroendocrine tumor, achieving F1 score of 0.81 (95% CI: [0.79,0.83]) for mitosis detection in cross-validation experiments. Through the MIDOG22 challenge, MDFS algorithm performance was evaluated across 10 unseen tumor types from different species and labs, including human melanoma, human astrocytoma, human bladder carcinoma, canine mammary carcinoma, canine mast cell tumor, human meningioma, human colon carcinoma, canine hemangiosarcoma, feline soft tissue sarcoma, and feline lymphoma, where our algorithm achieved overall F1 score of 0.764 (95% CI: [0.74,0.78]) outperforming all other algorithms and on-par with individual pathologists performance^95^.Mitosis detection on whole slide images We employed a WSI processing workflow^15^ in which TIAToolbox’s tissue segmentation model^102^ was used to delineate tissue regions. This model processes image tiles measuring 1024×1024 pixels at a resolution of 8 microns per pixel. Within the identified tissue regions, smaller tiles of 512×512 pixels were extracted at a higher resolution of 0.25 microns per pixel (corresponding to approximately 40× objective magnification), with a 50-pixel overlap between adjacent tiles. The MDFS algorithm subsequently identifies mitotic figures within these high-resolution patches. To integrate patch-level detections into a coherent WSI-level output, we applied a non-maximum suppression (NMS) strategy to address duplicate detections arising from overlapping regions. This approach ensured that redundant detections of the same mitotic figure across neighboring tiles were consolidated into a single representative detection, thereby producing accurate and spatially consistent WSI-level predictions.To account for the artifacts commonly found in real-world histology images (e.g., pen markings, stain residues, and debris), we extracted a collection of 14,000 small patches containing such artifacts from 30 WSIs sampled from the TCGA dataset. These patches were used alongside the MIDOG22 dataset^95^ to further refine the "Detecting Slow" stage of our MDFS pipeline. Subsequently, all diagnostic slides from TCGA were processed as previously described^15^. This process resulted in over 5 million mitotic candidates identified across 9,121 WSIs spanning 31 cancers, now publicly available at https://zenodo.org/records/14548480.

To further validate mitotic detection performance at the WSI level and ensure that our analysis was not influenced by excessive noisy detections, we conducted a review process. For this, 30 cases from each cancer type were randomly sampled, and the detections were validated by an experienced pathologist. Our pathologists found that the algorithmic predictions were generally accurate and acceptable for drawing biological conclusions. However, caution is warranted for some cases in ACC and MESO, where an increased number of false positives were observed in necrotic regions. Moreover, some cases in KICH exhibited a higher rate of false positives, primarily due to the substantial differences in nuclear appearance in this cohort compared to the images used for model training. We anticipate that future advances in mitosis detection algorithms will help mitigate these issues.

### Mitotic features

#### Hotspots detection and hotspot counts

In our study, we identified mitotic hotspot regions within WSIs using a deterministic algorithm based on mitoses detected by the MDFS model. A mitotic hotspot is classically defined as the area with the highest concentration of mitoses within a specified region of interest, often measured within a 2 or 3 mm² area^4,52^. To locate this region, we applied an overlapping window search across the WSI (stride of 0.3 mm), with each window covering an area of 3 mm² (1:1 ratio window). The algorithm determined the hotspot as the window containing the highest number of detected mitoses. The hotspot mitotic count (HSC) is defined as the mitotic count in the mitotic hotspot. We similarly define an atypical mitotic hotspot, which is identified using the same overlapping window search approach. However, instead of considering all detected mitoses, this search focuses exclusively on atypical mitoses which are detected using atypical mitosis detection model.

#### Mitotic Activity Network and Mitotic Topological Features

For each WSI, we construct a “mitotic activity network” by representing each detected mitosis as a node. To define edges, we consider the proximity of mitoses: any two nodes are connected if the spatial distance between their corresponding mitoses is lower than 400 µm (1/5 of the usual length of mitotic hotspot). This 400 µm radius was chosen heuristically by visualizing and comparing multiple WSIs across diverse tumor types to yield a biologically meaningful mitotic network that adapts to both low and high mitotic densities. By fixing the same threshold for all cases and cancers, the derived graph metrics remain directly comparable; although absolute feature values may vary with a different radius, it is their relative differences across samples that drive meaningful downstream analyses. By incorporating this distance-based criterion, we assume that mitoses within close proximity may interact or influence each other within the TME.

Once the mitotic network is formed, we apply social network analysis (SNA) techniques to derive a set of node-level measures to gain insights into the local connectivity and structural importance of each node within the mitotic network. The following measures^16,103^ are defined for each node *v*:

**Node Degree:** the *degree* of a node *v* in the network, denoted as ND(*v*), is the number of nodes directly connected to it. Formally, if we let *A* represent the adjacency matrix of the network, where *A*_*ij*_ = 1 if node *i* is connected to node *j* and 0 otherwise, then the degree of node *v* is defined as:

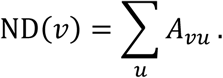

In the context of mitotic network, a mitotic figure (node) with a high degree is directly connected to many other mitoses within the 400 µm distance threshold. This indicates that the given mitosis is situated in a locally dense area of cellular activity, potentially reflecting a proliferatively active region of the tumor.

**Clustering Coefficient**: The *clustering coefficient* of a node *v*, denoted as CL(*v*), measures how interconnected its neighbors are by calculating the fraction of possible triangles through that node. Suppose deg(*v*) is the degree of node *v*, and *N*(*v*) is the set of its neighbors. The clustering coefficient is defined as:

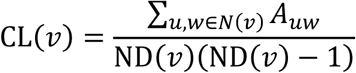

A high clustering coefficient for a mitosis indicates that its neighboring mitoses form a tightly knit cluster. In biological terms, this could suggest that the mitosis resides in a structurally coherent region of proliferative activity.

**Harmonic Centrality**: The *harmonic centrality* of a node *v*, denoted as HC(*v*), captures how easily a node can reach all other nodes in the network. If *d*(*v*, *u*) is the shortest path distance between node *v* and node *u*, then:

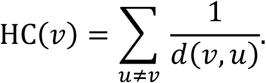

For a mitosis, a higher harmonic centrality means it is strategically positioned to access or influence many other mitoses with relatively few steps. This can be interpreted as the mitosis being located in a region where potential signaling or interactions with other proliferative cells can occur more efficiently.

#### Statistics of SNA measures as MTFs

Since the number of nodes and their corresponding SNA measures vary among different WSIs, simply using raw node-level values to compare mitotic activity of different WSIs is not possible. Instead, by extracting the statistics (mean and cv: coefficient of variation) of these measures into a set of fixed-size feature vector, we create a consistent and compact representation of the network’s topology. This approach enables meaningful comparisons and downstream analyses. The following table provides a list of extracted features, their meanings, and interpretations in the proposed mitotic network.

**Table 1:** Mitotic Topological Features (MTFs) and their interpretation in the proposed mitotic activity network.

| Feature | What is Being Measured? | Interpretation in the Mitotic Network |
| --- | --- | --- |
| mean(ND) | Average of node degrees across all mitoses. | Reflects the overall connectivity, indicating how dense mitotic interactions are on average. |
| cv(ND) | Coefficient of variation (standard deviation/mean) of node degrees. | Indicates the heterogeneity of connectivity; higher values suggest uneven distribution of mitotic connectivity, potentially indicating regions of differing mitotic activity. |
| mean(CL) | Average clustering coefficient of all nodes. | Shows the tendency of mitoses to form tightly interconnected groups, suggesting localized proliferation clusters. |
| mean(HC) | Average harmonic centrality of all nodes. | Reflects overall reachability within the network; higher values suggest that mitoses are generally well-positioned to influence or be influenced by others. |

#### Atypical Mitosis Detection (AMD)

For atypical mitosis detection, we trained EfficientNet-V2 Large^104^ model on a comprehensive dataset of histological image patches, including artifacts, mimickers, mitoses, and atypical mitoses.

To create such a dataset for atypical mitosis detection, we employed an AI-assisted labeling approach. First, we trained the initial AMD model using a combination of MIDOG22^95^ and TUPAC^105^ datasets for mitosis and mimicker categories, AMi-Br dataset^100^ for typical mitoses and atypical mitoses categories, along with our own artifact dataset. This initial dataset included approximately 9,500 image patches (64×64 pixels at 40x magnification).

Next, we sampled 6,000 representative patches from TCGA. To achieve this, small patches were extracted (64×64 pixels for 40x magnification WSIs or 32×32 pixels for 20x magnification WSIs which are then resized to 64×64 pixels). A pathology foundation model^106^ was used to generate feature embeddings from these patches. Using the K-Nearest Neighbors algorithm, the feature embeddings (patches) were clustered into 6 distinct groups (number of clusters were decided using the Elbow method). We then randomly sampled 1,000 patches from each cluster, yielding a diverse set of 6,000 candidates representative of the entire population.

To speed up the labeling process, we leveraged the initially trained AMD model to pre-classify the 6,000 sampled patches. These pre-classifications were subsequently reviewed and revised by three experienced pathologists. The finalized 6,000 mitotic patches were combined with the initial dataset, resulting in a dataset of 14,500 patches categorized into four classes: artifact, mimicker, mitosis, and atypical mitosis. A subset of this dataset is publicly available (https://zenodo.org/records/15390543).

Finally, we trained and evaluated the AMD model on the curated dataset. Using 3-fold cross-validation, we achieved a macro-averaged F1-score of 0.79 (SD0.03), a recall of 0.78 (SD 0.02), and a precision of 0.82 (SD 0.03) for classifying patches into the four categories. When applying the AMD model on the already detected mitotic candidates using MDFS, we only kept the candidates that were classified into either “mitosis” or “atypical mitosis” classes.

#### Atypical Mitosis Features (AMFs)

Using the sub-classification of atypical mitoses, we defined three measures to quantify atypical mitotic activity within the TME:

1. Atypical Mitosis in Atypical Hotspot (AMAH), which identifies the window with the highest count of atypical mitoses and reports that count.
2. Atypical Mitosis in Mitotic Hotspot (AMH), which represents the count of atypical mitoses within the identified mitotic hotspot
3. Atypical Mitosis Fraction in Whole Slide Image (AFW), calculated as the ratio of atypical mitoses to the total mitoses across the entire WSI.

### Survival analyses

We conducted pan-cancer survival analyses using recommended methodologies and four endpoints provided by Liu *et al.*^11^ to evaluate the prognostic utility of mitotic features. For each endpoint, we included only those cancer types with sufficient sample size and number of events to ensure reliable survival analysis and to prevent overestimation of the Concordance Index (C-index). In particular, for each endpoint, analyses were not performed for cohorts that have less than 100 cases or less than 10% event occurrence. All survival data used in these analyses were right-censored at 10 years (120 months) to ensure consistency across tumor types.

#### Permutation-based evaluation of prognostic value of mitotic features using Concordance Index

We assessed the prognostic discrimination of eight quantitative imaging features (HSC, mean(ND), cv(ND), mean(CL), mean(HC), AMH, AMAH, and AFW) across different tumor an event types by computing the C-Index between the raw feature values and survival data. For each cancer–feature pair, the observed C-Index was compared against a null distribution generated through 1000 permutations of the feature values, while keeping survival times and censoring statuses fixed. This permutation test provides a robust non-parametric framework that tests whether the observed concordance between a feature and outcome could arise by chance alone. The empirical two-sided p-value was calculated as the proportion of permuted C-Index values that were more extreme than the observed C-Index, relative to the null distribution centered at 0.5 (the expected C-Index under random association). To account for multiple comparisons across the large number of cancer–feature combinations, p-values were adjusted using the Benjamini–Hochberg procedure to control the false discovery rate. This method allowed us to identify which mitotic features show statistically reliable prognostic value across cancer types. Additionally, this framework facilitates cross-cancer comparison of prognostic performance for each feature.

#### Cross-validation for survival analysis

We employ a five-fold cross-validation approach for patient stratification across each cancer and endpoint combination^51^. For this, the dataset is partitioned into five folds based on event incidence to maintain a balanced representation of events in both training and test sets. Stratification can be performed using either raw feature values or partial hazard values obtained from univariate or multivariate Cox Proportional Hazards (CoxPH) model^107^.

In each fold, a decision threshold based on the median of the stratification variable (feature or hazard score) is calculated from the four training folds, and then the same threshold is used to dichotomize patients in the held-out test fold into high and low groups. This fold-wise stratification ensures that test data remain completely unseen during threshold selection, which is particularly important in survival studies with limited sample sizes and potential overfitting risks.

To construct cross-validated Kaplan–Meier (KM) curves, the test-set samples across all five folds are aggregated according to their assigned groups, and KM curves are generated for each group. This results in survival estimates based solely on out-of-sample predictions, allowing for an unbiased evaluation of the prognostic separation^51^.

#### Statistical significance testing in patient stratification

To evaluate the significance of survival differences between high-risk and low-risk groups, a permutation-based approach was applied^51^. In this statistical test, survival times were randomly shuffled with respect to covariates, and the entire cross-validation and KM curve construction process was repeated 1000 times. For each permutation, the log-rank statistic was computed for the resulting KM curves. The p-value was then determined as the proportion of permuted cases where the log-rank statistic was greater than or equal to the observed log-rank statistic from the original cross-validated KM curves. This provided a robust estimate of the likelihood that the observed separation between risk groups could have occurred by chance.

### Hot and Cold subgroups using Gaussian Mixture Models

In this paper, we categorized patients within each cancer type into Mitotic-Hot and Mitotic-Cold subgroups based on their mitotic activity (HSC), and into Immune-Hot and Immune-Cold subgroups based on CTLs expression. Instead of using a simple median of the desired feature within each cancer population to define the threshold for Hot and Cold populations, which would assume an approximately equal split of the population on either side of the threshold, we adopted a more biologically informed approach. Biological entities often exhibit variability that can be effectively modeled using normal (Gaussian) distributions.

Therefore, if the Hot and Cold populations each follow a Gaussian distribution, we can hypothesize that the observed distribution of the desired features (HSC or CD8) in a cancer type is a mixture of two Gaussian distributions.

To uncover these underlying distributions within the combined data, we employed the Gaussian Mixture Model (GMM). GMM is a probabilistic framework that models observed data as a combination of multiple Gaussian distributions. By fitting a GMM to the data, we estimated the parameters (means and variances) of the underlying distributions.

#### Finding the Hot-Cold intersection

By estimating the underlaying Gaussian distributions, we were able to identify a more meaningful method to classify cases into Hot or Cold populations. The intersection point between two Gaussian distributions is the threshold where their probability density functions (PDFs) are equal. At this point, the likelihood of a sample belonging to either subgroup is identical, making it a natural boundary to separate the subgroups.

For two Gaussian distributions 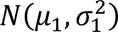 and 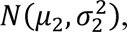 their PDFs are defined as:

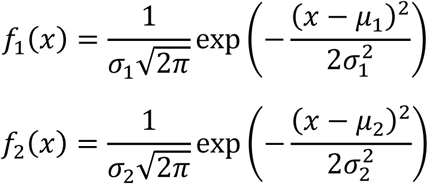

To find the intersection, we set *f*_1_(*x*) = *f*_2_(*x*):

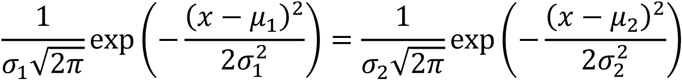

Taking the natural logarithm of both sides simplifies the equation:

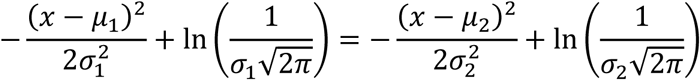

Rearranging terms, we obtain:

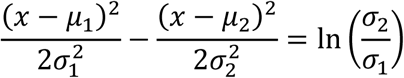

Expanding the squares and collecting terms yields a quadratic equation:

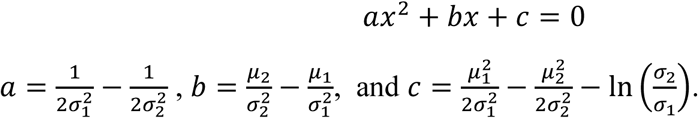

where: 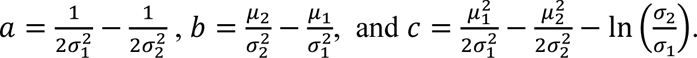

The roots of this quadratic equation are calculated using the quadratic formula:

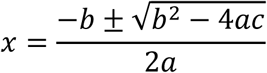

We select the root *x* that lies between *μ*_1_and *μ*_2_, as it represents the intersection point of the two distributions. This intersection point is calculated for mitotic and immune activity in each cancer separately to categorize the patients into Hot or Cold subgroups.

### Genomic analyses

#### Cross-validated canonical correlation

To investigate the linear relationship between the proposed MTFs and the gene set associated with mitotic activity as identified in the literature, we employed canonical correlation analysis (CCA). CCA is a statistical method that identifies and quantifies the relationships between two sets of variables. Mathematically, it works by finding linear combinations of the variables in each set such that the correlation between the resulting canonical components is maximized. For two datasets, **X** and **Y**, CCA seeks vectors **a** and **b** that maximize the correlation ρ = corr(**Xa**, **Yb**).

To ensure robustness when applying CCA to our dataset, we implemented a five-fold cross-validation approach. Specifically, the dataset was randomly divided into five folds. For each iteration, the CCA model was trained using four folds (training set) and tested on the remaining hold-out fold. The testing involved measuring the correlation between the first canonical component of the MTFs and the first canonical component of mitotic gene expression. This process was repeated across all five folds, and the average correlation value across the test folds was reported as the final measure of the linear relationship.

#### Quantifying mutation–phenotype associations via AUC and permutation testing

To assess associations between somatic mutation status and spatial features of mitotic activity, we evaluated the relationship between binary mutation labels (mutated vs. wild-type) and continuous MTF values using an area under the receiver operating characteristic curve (AUC)–based approach^22^. AUC was chosen because it is suitable for quantifying the strength and direction of association between a binary variable and a continuous phenotype.

Furthermore, AUC can also reflect the potential predictive value of the features if used in models aimed at classifying mutation status. For each gene-feature pair, the AUC was computed to evaluate whether samples harboring a specific mutation systematically exhibit higher or lower MTF values than wild-type samples. To aid interpretation and to center the metric around zero, we linearly transformed the AUC scores from their native range of [0.5, 1] (or [0, 0.5] if reversed) to a symmetric range of [–1, 1] using the formula: Transformed AUC=2×(AUC−0.5). Under this transformation, a value of 0 indicates no association, positive values indicate that mutation-positive cases tend to have higher MTF values than wild-type, and negative values indicate the opposite.

To assess statistical significance, we performed a permutation-based test for each gene-feature pair. Specifically, we first calculated the observed AUC for the real mutation status labels. Then, we shuffled the mutation labels across samples 1,000 times while preserving the MTF values, recalculating the AUC for each permuted dataset. The empirical p-value was computed as the proportion of permutations where the permuted AUC exceeded the observed AUC in the direction of effect (i.e., greater than the observed AUC if positive, or less than the observed AUC if negative). Finally, p-values were adjusted for multiple hypothesis testing using the Benjamini–Hochberg procedure to control the false discovery rate. Associations with an q-value < 0.05 were considered statistically significant.

#### Gene Set Enrichment Analysis

For all gene expression comparisons, raw gene count matrices and corresponding phenotype data were used to perform differential gene expression analysis and gene set enrichment analysis (GSEA)^34^. Sample-specific expression count data were first filtered to retain genes with sufficient expression (minimum total counts ≥ 100 across samples) and samples with matched phenotype information. Gene-level differential expression was assessed using the Python implementation of the DESeq2 method^108^ via the “PyDESeq2” package^109^. Comparisons were made between two conditions (e.g., Mitotic-Hot vs. Mitotic-Cold), with model fitting and statistical testing based on negative binomial generalized linear models. Genes with an q-value < 0.001 and |log_2_(*fold change*)| > 1 were considered significantly differentially expressed.

For GSEA, genes were ranked by the DESeq2 test statistic, and pre-ranked enrichment analysis was performed using the GSEAPY package^110^. The goal of GSEA is to determine whether predefined gene sets (e.g., biological pathways) are significantly overrepresented at the top or bottom of the ranked list, indicating coordinated up- or down-regulation across a biological process^34^. Pathways were considered significantly enriched at FDR q-value < 0.01. Analyses were performed against multiple gene set libraries, including Human MSigDB Hallmark 2020, KEGG 2021 Human, and Reactome 2022, using a fixed random seed to ensure reproducibility.

### Statistical tests

Multiple statistical approaches were employed throughout the study to assess associations, compare distributions, and evaluate significance. For specific permutation-based analyses (such as evaluating the statistical significance of AUC scores, C-Index, and cross-validated KM survival curves) nonparametric permutation testing was used. These tests are described in detail in the relevant sections of the Material and Methods section.

For correlation analyses, standard two-sided p-values were reported alongside Spearman or Pearson correlation coefficients, depending on the assumptions about monotonicity and linearity of the relationship, respectively. In multivariable survival analysis models, statistical significance of survival differences between groups was assessed using log-rank tests.

To compare the distributions of continuous variables across two groups, we used the two-sided Mann–Whitney U test (also known as the Wilcoxon rank-sum test). This nonparametric test was chosen because it does not assume normally distributed data and remains valid even when the two groups have different underlying distributions or sample sizes.

All reported p-values were adjusted for multiple hypothesis testing using the Benjamini– Hochberg FDR correction.

## Supplementary Tables

Excel files for supplemental tables can be found in this link: https://zenodo.org/records/14793678.

Table S2: Excel file containing list of PACMAN genes and their correlations with MTFs

Table S3: Excel file containing full results of measuring association of gene mutations with MTFs.

Table S4: Excel file containing full results of measuring association of methylation CpG with MTFs.

Table S5: Excel file containing full results of ANOVA analysis of MTFs association with CNV.

Table S8: Excel file containing full results of multivariate survival analysis using AFW and HSC.

Table S9: Excel file containing full results of multivariate survival analysis using AMAH and HSC.

Table S10: Excel file containing full results of multivariate survival analysis using AMH and HSC.

Table S11: Excel file containing full results of GSEA between AFW-High and AFW-Low subgroups.

## Supplementary Figures

**Figure S1:**
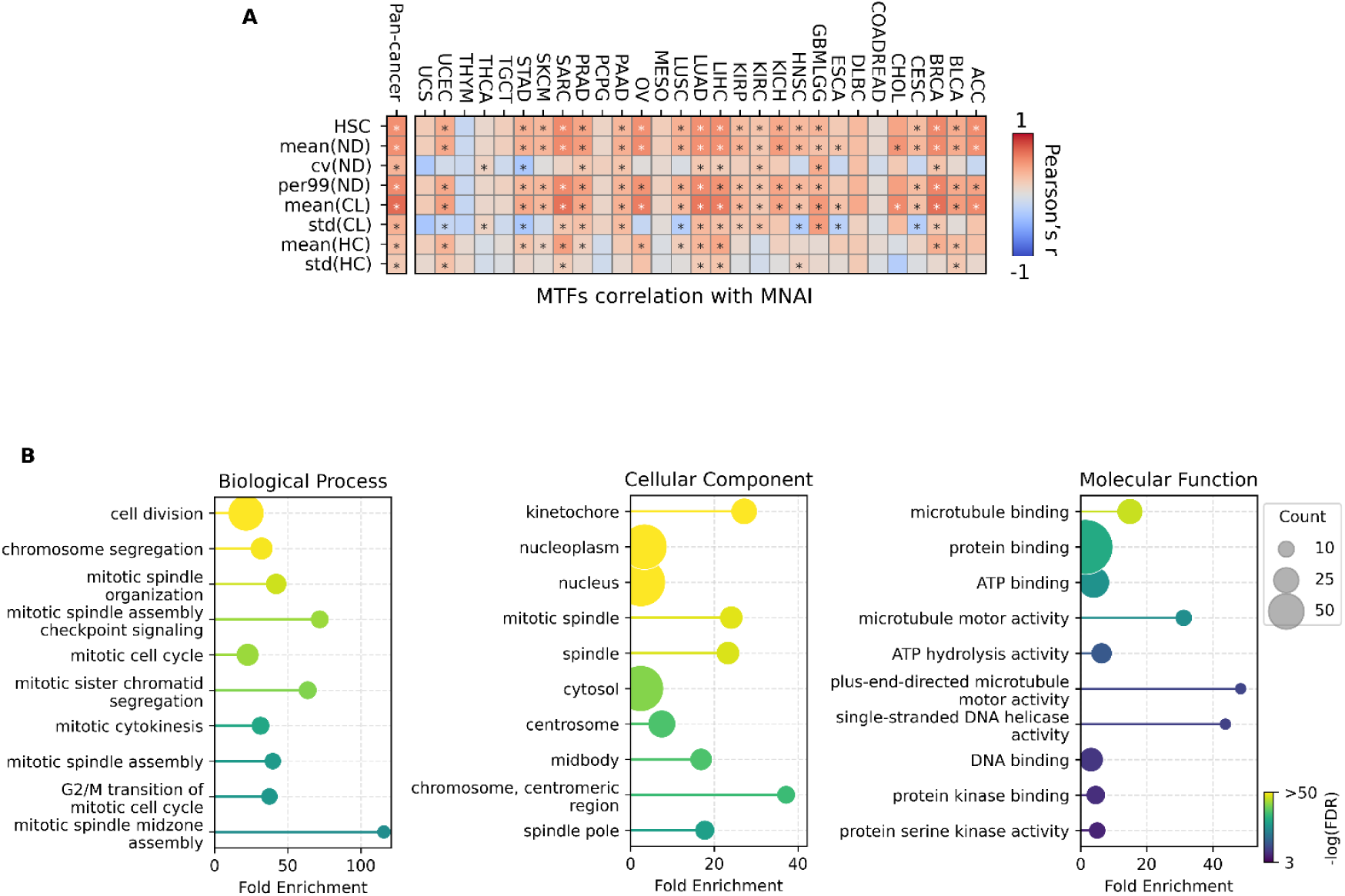
Validating MTFs as mitotic activity descriptors. (A) Heatmap of Pearson’s correlation (*r*) between MTFs and MNAI. Asterisks indicate significant correlations (q-values<0.05). (B) Gene ontology analysis using the PACMAN gene set.

**Figure S2:**
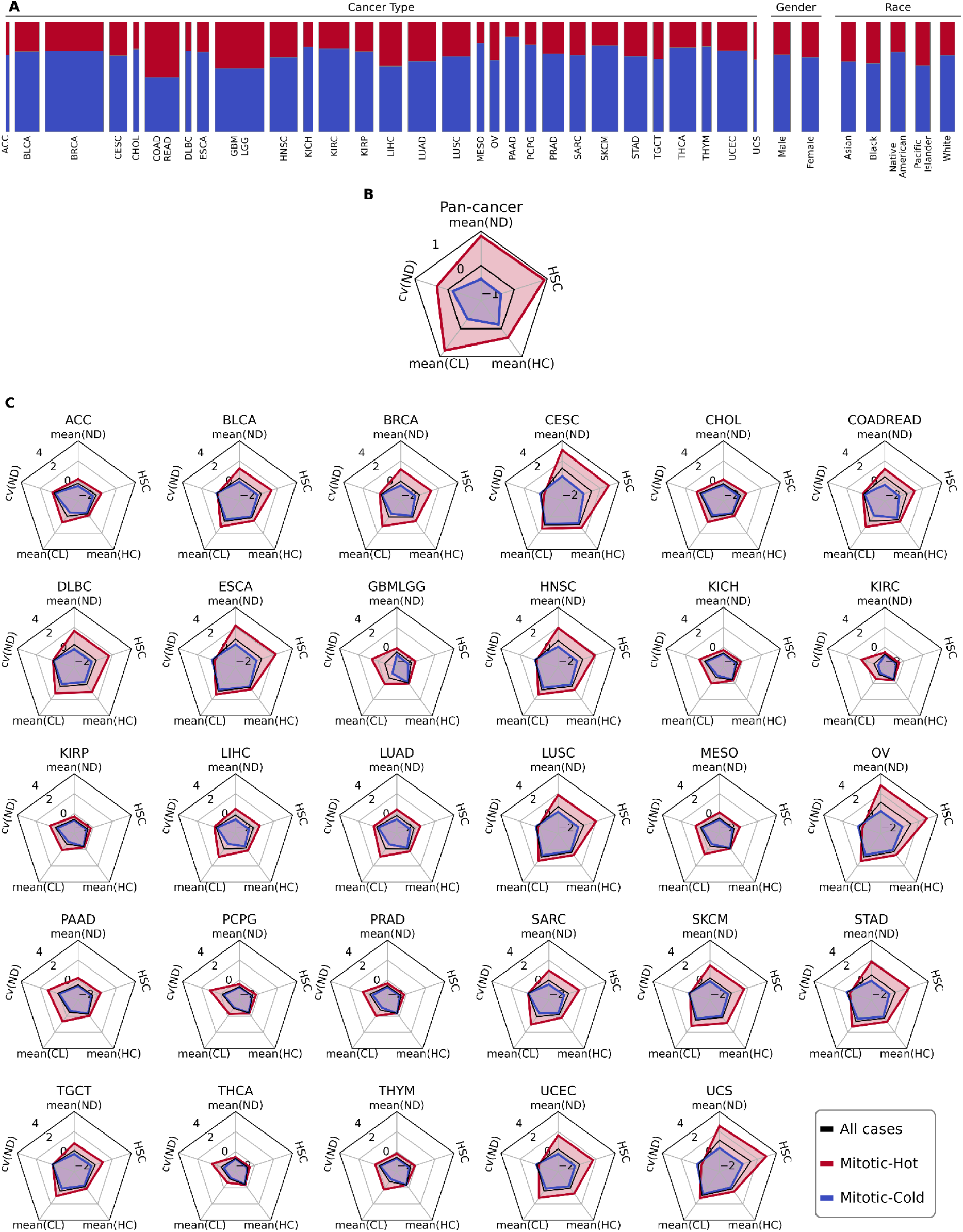
Statistics of Mitotic-Hot and Mitotic-Cold subgroups. (A) Proportion of Mitotic-Hot to Mitotic-Cold groups in each cancer type, where the width of each bar represents the population of that cohort. Radar plots showing distributions of MTFs over Mitotic-Hot and Mitotic-Cold subgroups on pan-cancer level (B) and cancer-specific scenario (C).

**Figure S3:**
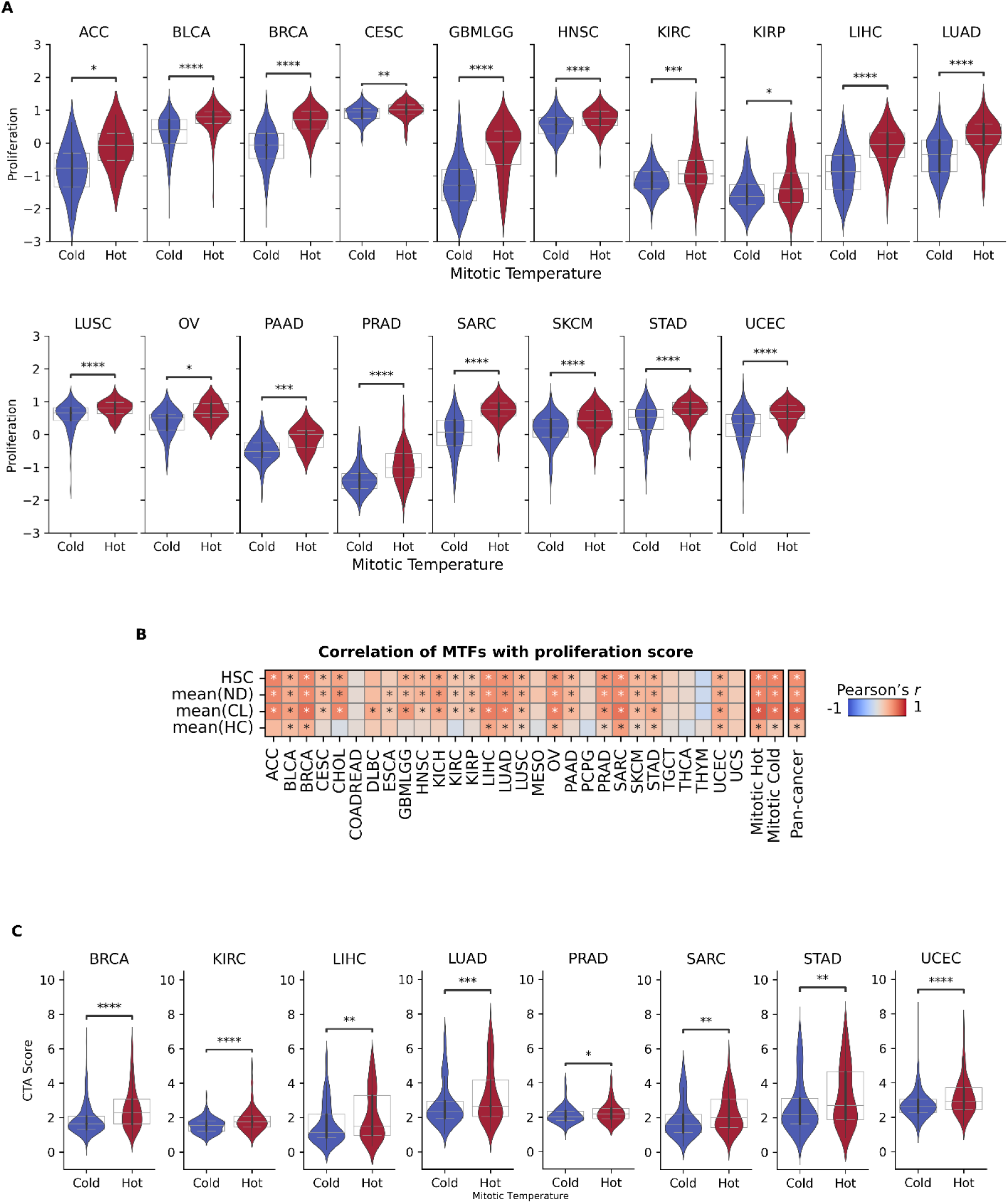
Tumor characteristics over Mitotic-Hot and Mitotic-Cold subgroups. (A) Violin plots showing the distribution of Proliferation score over mitotic-hot and cold groups. (B) Correlation of MTFs with proliferation score driven from genomic information. Asterisks denote Bonferroni-adjusted p-values <0.05. (C) Violin plots showing the distribution of CTA scores over Mitotic-Hot and Mitotic-Cold groups. In (A) and (C), Asterisk indicate statistical significance based on Bonferroni-adjusted p-values resulted from Mann-Whitney U test (*: p-value<0.01, **: p-value<0.001, ***: p-value<0.0001, ****: p-value<0.00001).

**Figure S4:**
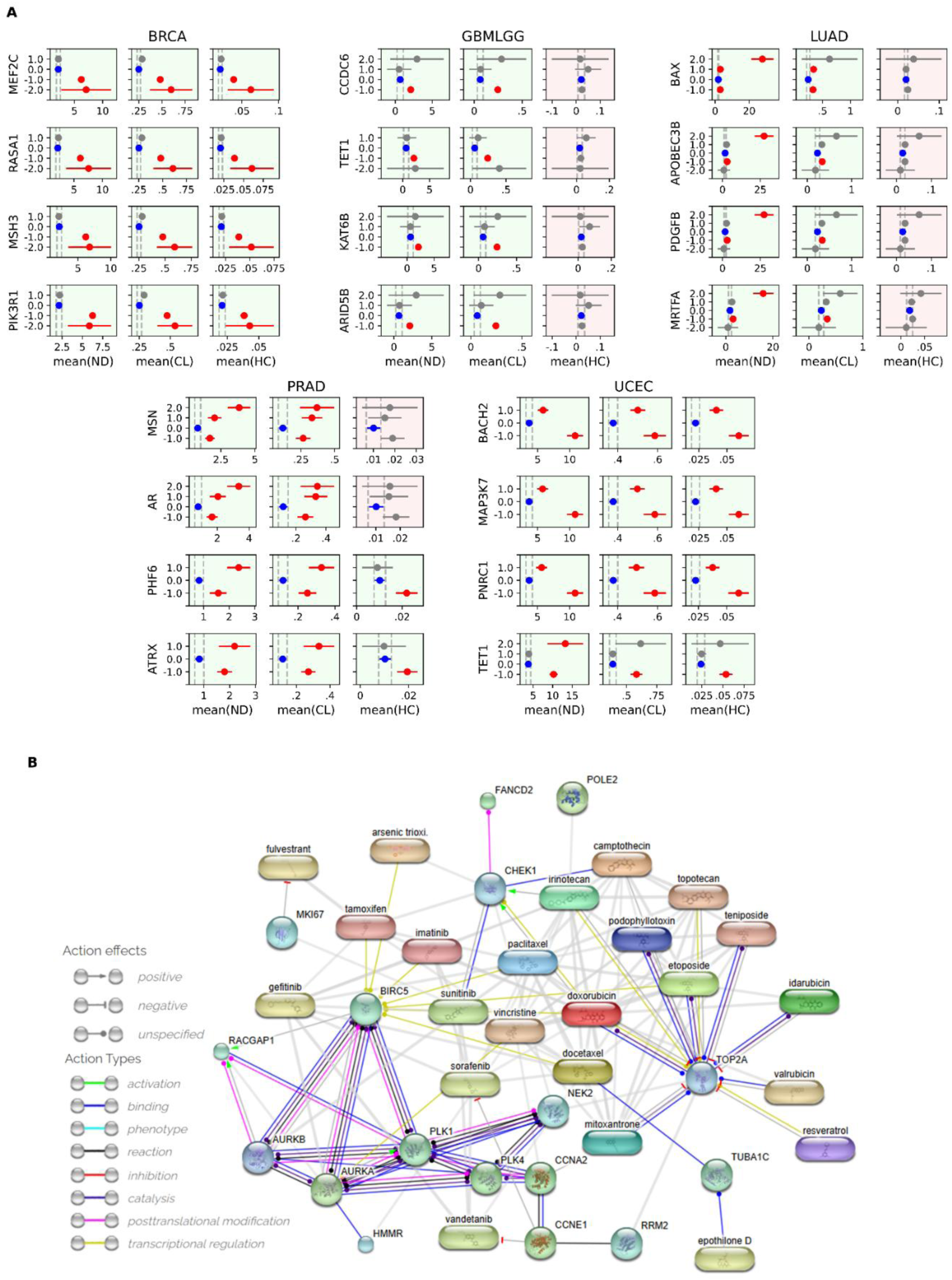
(A) Tukey’s post-hoc analyses to visualize MTFs’ ranges over different states of CNV in genes with significant differences identified through ANOVA analyses (related to Figure 3B). (B) Protein-Protein-Drug network based on PACMAN gene set.

**Figure S5:**
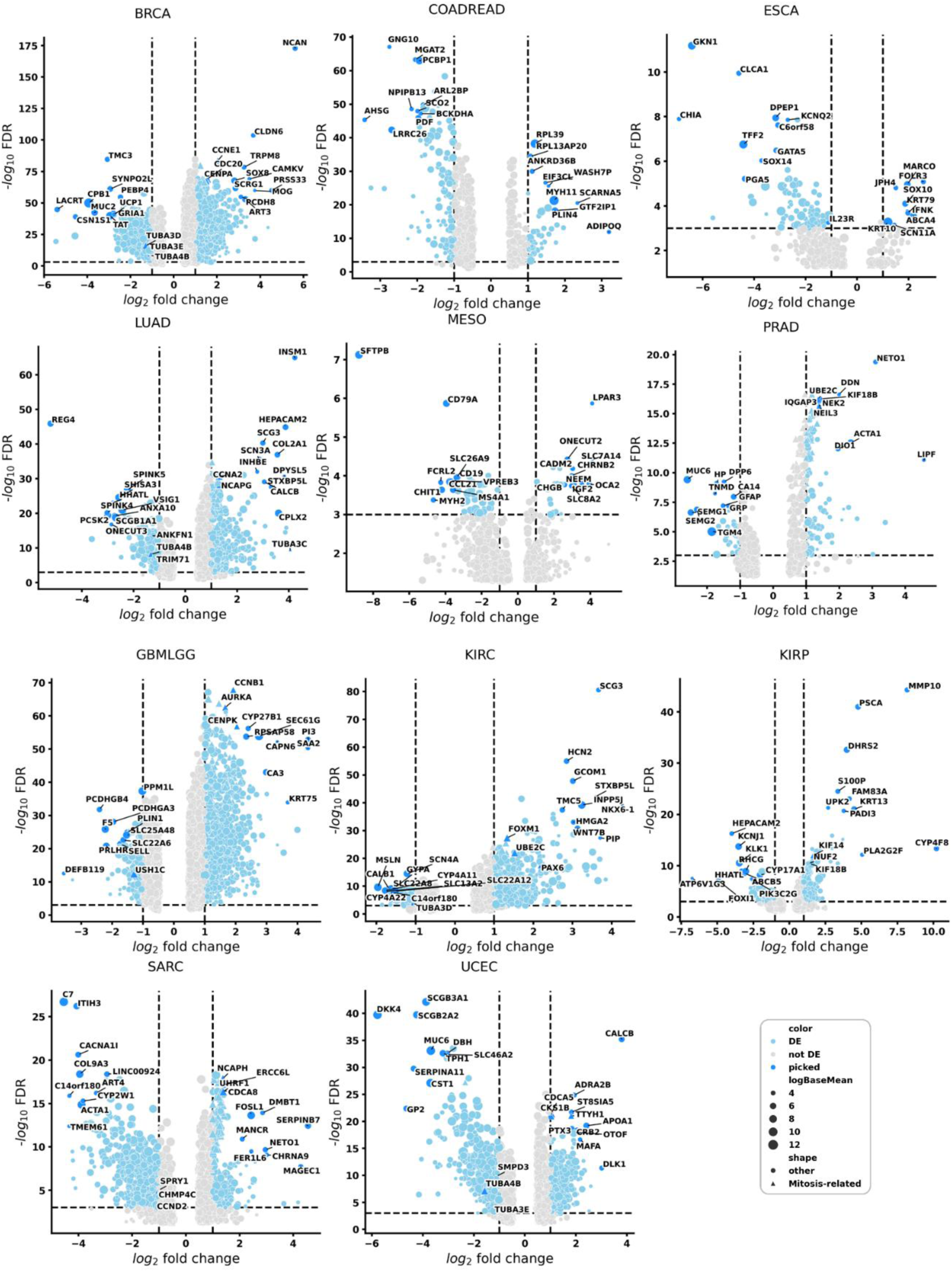
Volcano plot of differentially expressed genes between Mitotic-Hot vs Mitotic-Cold subgroups (related to Figure 3C).

**Figure S6:**
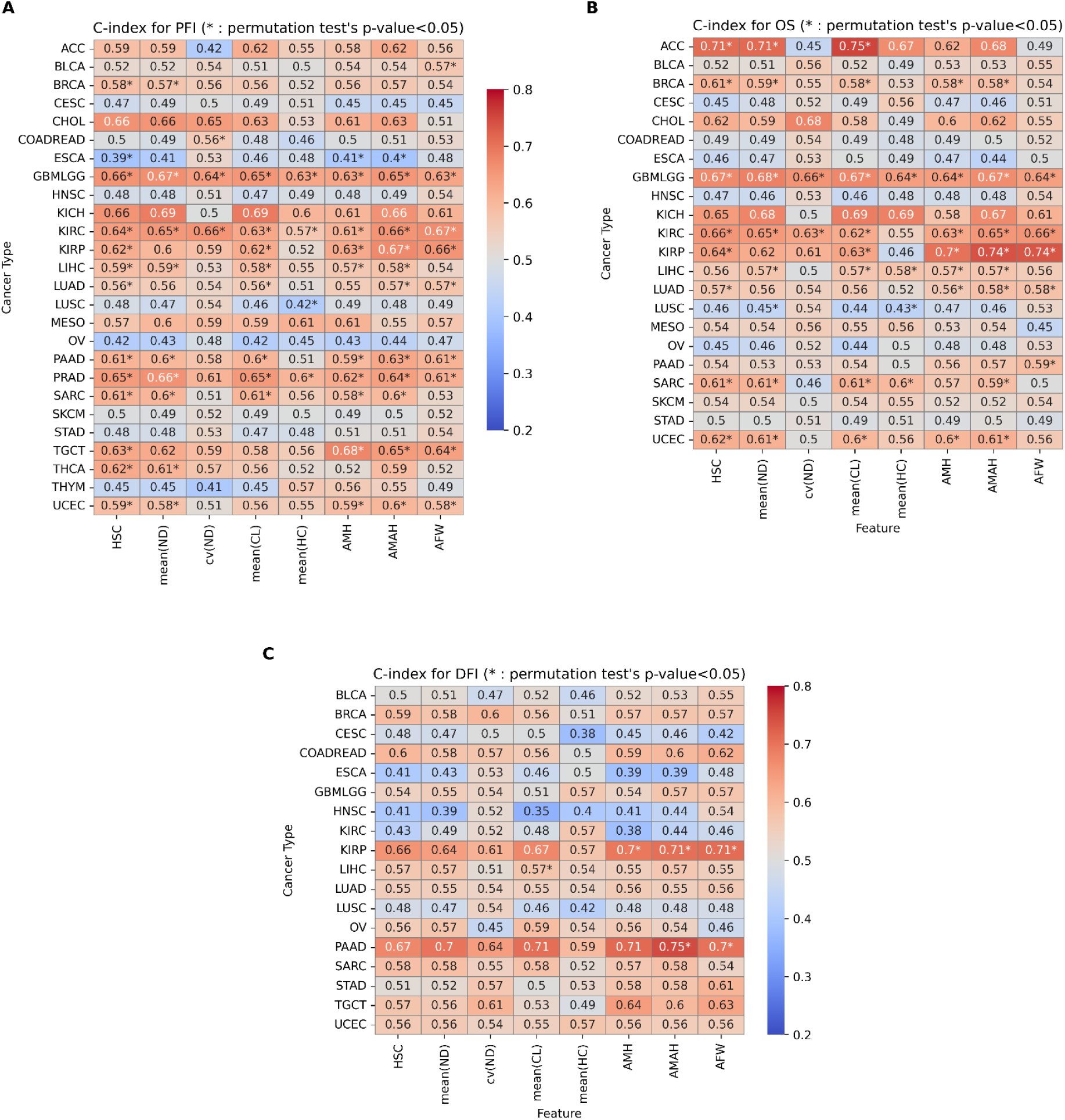
C-indices of MTFs and AMFs for (A) PFI, (B) OS, and (C) DFI ranking.

**Figure S7:**
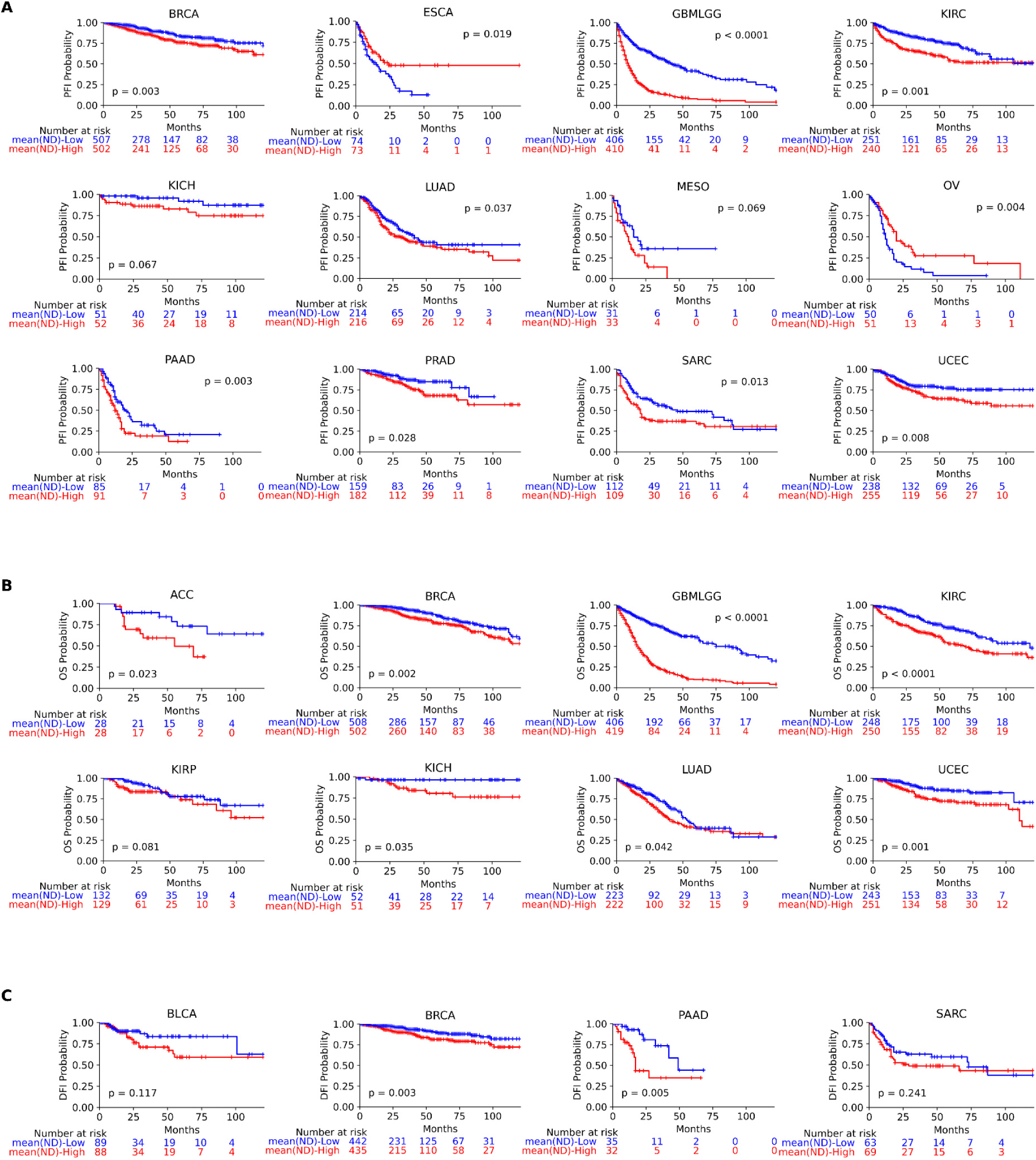
Cross-validated KM curves for mean(ND)-Low and mean(ND)-High patients. based on (A) PFI, (B) OS, and (C) DFI probabilities. Here, p-values show statistical significance through permutation tests.

**Figure S8:**
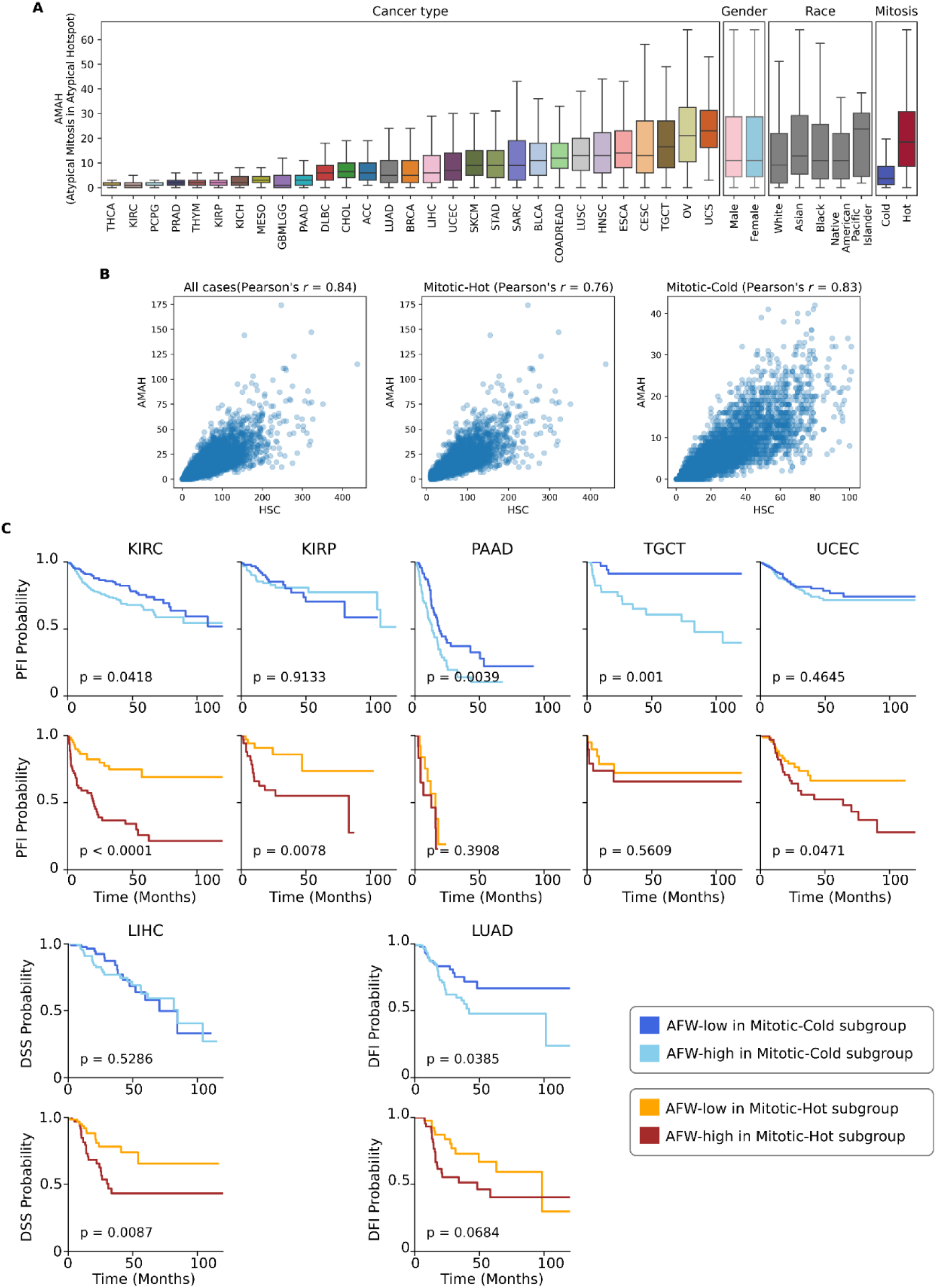
Distribution atypical mitoses and their prognostic value across cancers. (A) Box plots showing distribution of atypical mitotic activity (through AMAH) over different cancer types, genders, races, and mitotic temperatures. (B) Correlation of AMAH with HSC across all cases, Mitotic-Hot, and Mitotic-Cold subgroups. (C) (F) KM curves showing AFW-High and AFW-Low patient stratification (based on the median AFW) in Mitotic-Hot and Mitotic-Cold subgroups, separately. Here, p-vales report statistical significance of stratification through log-rank test.

